# Social health and subsequent cognitive functioning in people aged 50 years and above: examining the mediating roles of depressive symptoms and inflammatory biomarkers

**DOI:** 10.1101/2023.11.02.23297985

**Authors:** Jean Stafford, Serhiy Dekhtyar, Anna-Karin Welmer, Davide L Vetrano, Giulia Grande, Anna Marseglia, Vanessa G Moulton, Rosie Mansfield, Yiwen Liu, Ke Ning, Karin Wolf-Ostermann, Henry Brodaty, Suraj Samtani, Mohammad Arfan Ikram, René Melis, Joanna Rymaszewska, Dorota Szcześniak, Giorgio Di Gessa, Marcus Richards, Daniel Davis, Praveetha Patalay, Jane Maddock, the SHARED Consortium

## Abstract

**Background:** Social health markers, including marital status, contact frequency, network size, and social support, have shown associations with cognition. However, the underlying mechanisms remain poorly understood. We investigated whether depressive symptoms and inflammation mediated associations between social health and subsequent cognition.

**Methods:** In the English Longitudinal Study of Ageing (ELSA; n=7,136; aged 50+), we used four-way decomposition to examine to what extent depressive symptoms, C-reactive protein (CRP) and fibrinogen (assessed at an intermediate time point) mediated associations between social health and subsequent standardised cognition (verbal fluency, delayed and immediate recall) including cognitive change, with slopes derived from multilevel models (ELSA: 12-year slope). We examined whether findings replicated in the Swedish National Study of Aging and Care in Kungsholmen (SNAC-K; n=2,846; aged 60+; 6-year slope).

**Findings:** We found indirect effects via depressive symptoms of network size, positive support and less negative support on subsequent verbal fluency, and positive support on subsequent immediate recall (pure indirect effect (PIE)=0.002 [0.000-0.003]). The positive support-verbal fluency mediation finding replicated in SNAC-K. Depressive symptoms partially mediated associations between less negative support and slower immediate (PIE=0.001 [0.000-0.002]) and delayed recall decline (PIE=0.001 [0.000-0.002]), and between positive support and slower immediate recall decline (PIE=0.001, [0.000-0.001]), which replicated in SNAC-K. We did not observe mediation by inflammatory biomarkers.

**Interpretation:** Findings provide new insights into mechanisms linking social health with cognition, suggesting that associations between cognition and interactional aspects of social health in particular, such as social support, are partly underpinned by depressive symptoms.

## Introduction

Social health, an umbrella term encompassing aspects of social relationships from the individual level to wider socio-cultural factors,^1^ is increasingly recognised as a crucial component of health across the life course, especially in older people. Multiple aspects of social health, such as marital status, social isolation and loneliness, have been linked with diverse health-related outcomes, including mental health,^2^ cardiovascular disease,^3^ and mortality.^4^

Distinct markers of social health, including social isolation, loneliness and marital status, have also been associated with subsequent cognitive decline and dementia.^5–8^ Our previous cross-cohort research revealed that positive aspects of social connections were associated with lower risk of dementia,^9^ and subsequent cognitive functioning, including cognitive change, in those without dementia.^10^

However, substantial gaps remain in understanding the mechanisms underpinning the relationships between social health and cognition. Several pathways have been posited to underlie these relationships, including depressive symptoms, given the potential for social relationships to provide feelings of security and emotional support, and to buffer against stress.^11^ In line with this, social disconnectedness and perceived isolation have shown bidirectional relationships with depression and anxiety among older people,^12,13^ and psychiatric symptoms are, in turn, associated with increased risk of cognitive decline and dementia.^14–16^ Both late-life depression and social isolation were identified as modifiable risk factors in the Lancet commission on dementia prevention, intervention and care.^17^

Inflammation has also been proposed as a pathway which could underlie associations between social health and cognition, given that aspects of social health such as loneliness and social isolation have shown associations with inflammatory biomarkers, including C-Reactive protein (CRP) and interleukin-6.^18^ Inflammatory biomarkers have also been linked with increased risk of cognitive decline and dementia.^19^ However, there is limited evidence to date about the possible mediating roles of depressive symptoms or inflammation in the relationships between social health and cognitive functioning.

While several previous studies indicated a possible mediating role of depressive symptoms,^20–24^ these studies were cross-sectional or had relatively short follow-up periods (between 4-10 years). Studies examined varied social health markers, including loneliness,^20,22^ friendship,^21^ cohabitation,^23^ and social support,^24^ and mainly focused on global cognition, rather than individual cognitive domains. Fewer studies examined inflammation as a mediator, with two studies finding no evidence of mediation in relation to loneliness,^20,25^ and one finding a mediating role in the association between social isolation and cognition in men only.^26^ These studies were cross-sectional or had relatively short follow-up periods, did not examine cognitive trajectories, and one combined inflammatory biomarkers with other physiological markers.^20^ Further research is required to investigate the mediating role of depressive symptoms and inflammation in representative samples with longer follow-up periods, including examination of cognitive trajectories, and investigating a range of social health markers and individual cognitive domains.

To address these gaps in knowledge, using the English Longitudinal Study of Ageing (ELSA), we investigated whether and to what extent depressive symptoms and inflammation mediated associations between social health and subsequent cognitive functioning, including cognitive trajectories. We examined whether findings replicated in the Swedish National Study of Aging and Care in Kungsholmen (SNAC-K). We focused on both structural aspects of social health, including marital and cohabitation status, network size and frequency of contact, and interactional aspects such as perceived social support.

## Methods

### Design

We conducted primary analysis in ELSA and replication analysis in SNAC-K. Examining replication provides insight into whether findings are similar across studies and settings, thereby enhancing confidence in the replicability of findings, or whether findings differ, suggesting a more cautious interpretation may be needed.^27^

### Primary sample (ELSA)

#### Data source and participants

ELSA is an ongoing nationally representative survey of participants aged 50 years and older living in private households in England. Participants were initially recruited in 2002-2003 (individual response rate 67%), with eight subsequent waves taking place every two years. Our analytic sample (n=7,136) included participants aged 50 years and older without recorded dementia at baseline or the intermediate time point in which mediators were assessed (Figure S1), with recorded information on at least one social health marker and potential mediator, and three assessments of the same cognitive outcome between waves 3-9 (with one assessment in wave 3).

#### Measures

Full details of questions and coding for exposures, outcomes, mediators and covariates is provided in the Supplementary Material.

#### Social health exposures (baseline)

Structural social health markers included marital/cohabitation status, network size based on the number of children, family, and friends with whom participants reported having a close relationship (capped at 30 people), and frequency of contact with children, friends and family members in person, by phone and/or through writing or email (never to every few months; once or twice a month; at least weekly). Interactional social health markers included perceived positive social support from partner, children, friends and other family, a numeric variable where higher scores indicated more support averaged across relationship types. Perceived negative support was assessed across the same relationship types, with higher scores indicating less negative support.

#### Potential mediators (intermediate time point)

Mediators were assessed at an intermediate time point between exposures and outcomes, including depressive symptoms measured using the 8-item Center for Epidemiologic Studies Depression Scale (CES-D),^28^ and inflammation (CRP (mg/L) and fibrinogen (g/L), with blood concentrations obtained from fasted blood samples. We log-transformed CRP for analysis (details in Supplementary Material).

#### Cognitive outcomes

We examined cognitive outcomes at a single time point following mediator measurement (wave 3), and cognitive change between waves 3-9. Cognitive outcomes included verbal fluency and immediate recall in ELSA and SNAC-K, and delayed recall, which was only available in ELSA (tasks described in Supplementary Material). We standardized each test across time points and within studies on a common standard deviation-based scale with a mean of zero and a standard deviation (SD) of one.

#### Covariates (baseline)

Covariates included age in years, sex (male; female), educational attainment (lower than secondary or none; secondary; higher education), occupational class (manual; non-manual), total non-pension household wealth quintiles, vascular-related health conditions (none; 1+), basic and instrumental activities of daily living (none; 1+), smoking status (never; ex-smoker; current smoker), physical activity (inactive; moderately active; highly active), alcohol consumption (not at all in the last year; monthly or less; around weekly; almost daily), baseline cognition and depressive symptoms, as described above. Directed acyclic graphs are presented in Figures S2 and S3.

### Replication sample (SNAC-K)

SNAC-K is an ongoing population-based longitudinal study which began in 2001 and 2004, with 5,111 people aged ≥60 years living at home or in institutions in Kungsholmen (central Stockholm). SNAC-K began with participants invited to participate in the baseline assessment. 2,848 participants responded and took part in cognitive tests and psychological testing. Younger age cohorts (60, 66, and 72 years) were followed every 6 years (2007-2010 and 2013-2016; the 72 age-cohort was further assessed in 2010-2013) and older age cohorts (≥80 years) every 3 years (2004-2007, 2007-2010, 2010-2013, and 2013-2016).

We included data from baseline, 3-year, 6-year, 9-year, and 12-year follow-up examinations. We used comparable measures across ELSA and SNAC-K wherever possible, although not all variables included in ELSA were available in SNAC-K, hence examination of replication is limited to available variables. Full information about exposures, mediators, cognitive outcomes and covariates is provided in the Supplementary Material. Social health markers were assessed at baseline and included marital/cohabitation status, network size, contact frequency and positive social support. Potential mediators, depressive symptoms and CRP, were assessed at an intermediate time point (6-year follow-up). Cognitive outcomes, verbal fluency and immediate recall, were assessed at the 12-year follow-up, and cognitive change was assessed between the 6-year and 12-year follow-up.

### Analysis

We examined the mediating roles of depressive symptoms and inflammation using a causal mediation approach based on the counterfactual framework.^29,30^ Before completing mediation analysis, we examined associations between exposures, mediators and outcomes. We only tested for mediation where we observed associations between a) a given exposure and mediator, and b) a given mediator and outcome.^31^ We did not require an overall exposure-outcome association, as recent methodological developments have shown that mediation can be examined where there is theoretical interest even when there is no significant total effect.^32^

First, we examined associations between social health markers and potential mediators using linear regression models. Second, we tested associations of social health exposures and mediators with subsequent cognition at a single time point, and cognitive trajectories derived from mixed-effects multilevel models. The date of individual interview (month/year) at each wave was used as the time metric, centered to date at baseline. To enable inclusion in mediation analyses, we extracted individual-level predicted slopes as indicators of cognitive change over time, expressed as standardized change per decade. Associations between exposures, mediators and extracted cognitive trajectories were tested using linear regression models before completing mediation analysis.

Next, we completed four-way decomposition of the total effect (TE) into the controlled direct effect (CDE), the reference interaction effect (INT_ref_), the mediated interaction effect (INT_med_) and the pure indirect effect (PIE).^33^ The CDE refers to the portion of the TE of social health markers on cognitive outcomes due to pathways not involving depressive symptoms or inflammatory markers (neither mediation nor interaction). INTref is the portion of the TE due to interaction, but not mediation, and INTmed is the portion due to interaction and mediation. PIE is the effect only due to mediation that does not involve interaction. Replication across studies was examined by qualitative comparison of effect estimates and 95% confidence intervals. We completed sensitivity analysis to examine associations between exposures and depression as a binary outcome indicating high levels of depressive symptoms using logistic regression, and between binary depressive symptoms and cognition. We also examined associations between exposures and CRP, and between CRP and cognition, after excluding those with levels higher than 10mg/L, which may indicate an acute infection or serious illness.

## Results

### Primary sample - ELSA

Descriptive information is provided in Table 1 and Table S1. The analytical sample included 7,136 participants (55.5% female, mean baseline age: 63.8; standard deviation (SD): 9.4; Table 1). Before testing for mediation, we examined associations between social health and cognition (Tables 2-3). After partial adjustment, social health markers were associated with cognition across most domains, with attenuation after full adjustment, except for less negative support, which remained associated with higher verbal fluency (β=0.02, 95%CI: 0.00 – 0.03). After full adjustment, higher contact frequency and less negative support (β=0.005, 95%CI: 0.000 – 0.009) were associated with slower immediate recall decline, and positive support with slower delayed recall decline (β=0.007, 95%CI: 0.002 – 0.012).

**Table 1.**
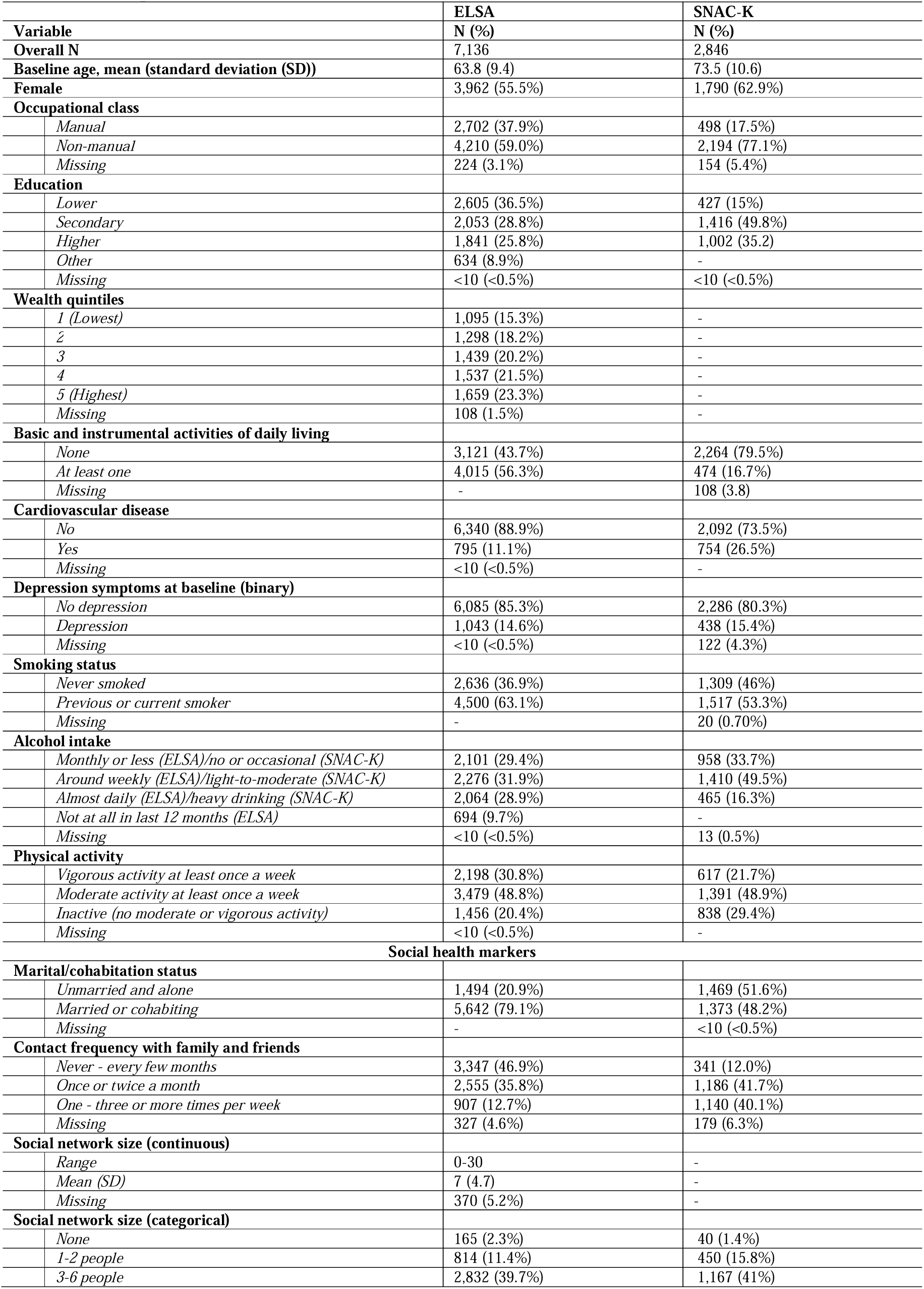
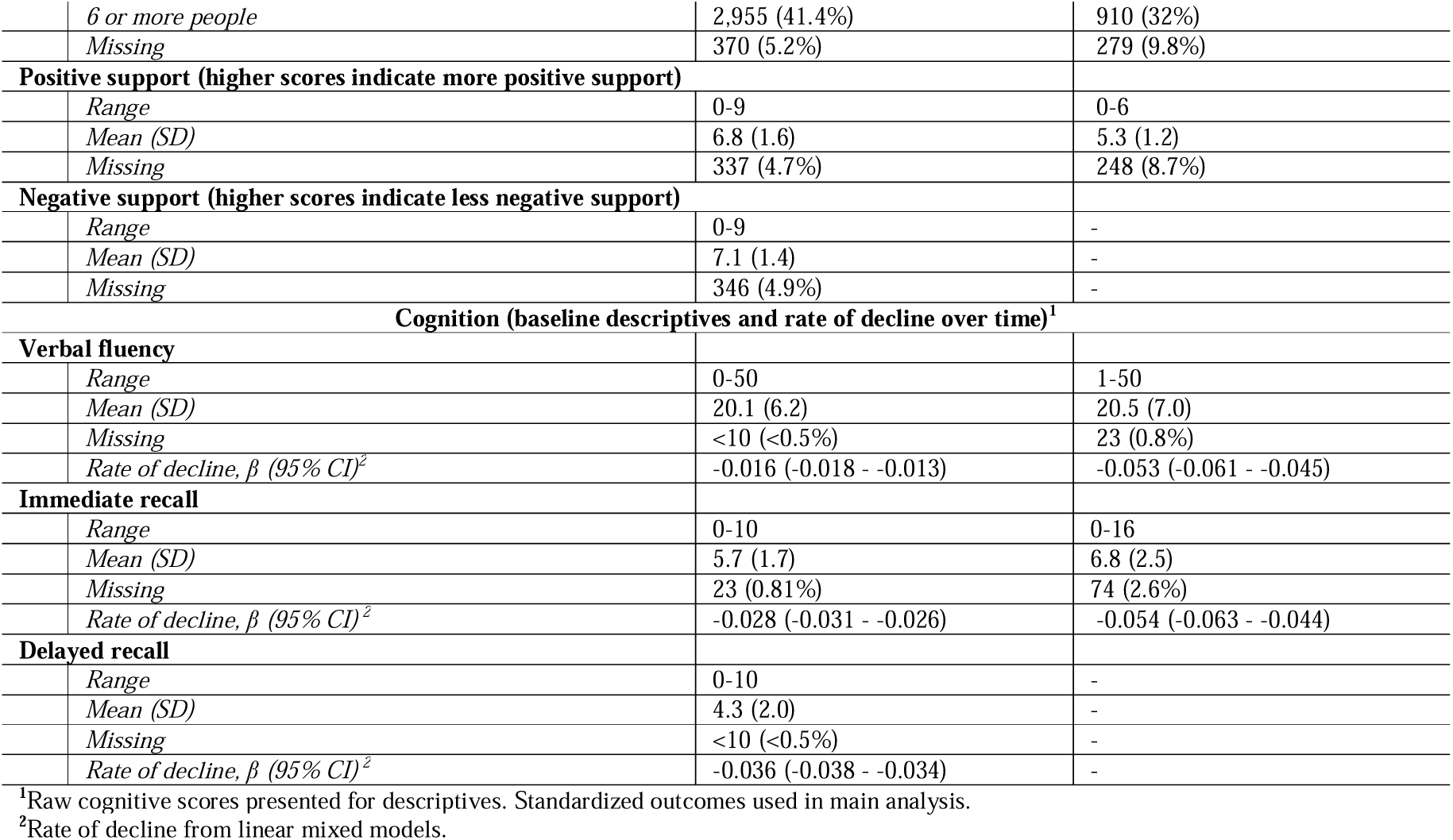
Descriptive statistics for social health markers and covariates.

|  | ELSA | SNAC-K |
| --- | --- | --- |
| Variable | N (%) | N (%) |
| **Overall N** | 7,136 | 2,846 |
| **Baseline age, mean (standard deviation (SD))** | 63.8 (9.4) | 73.5 (10.6) |
| **Female** | 3,962 (55.5%) | 1,790 (62.9%) |
| **Occupational class** |  |  |
| *Manual* | 2,702 (37.9%) | 498 (17.5%) |
| *Non-manual* | 4,210 (59.0%) | 2,194 (77.1%) |
| *Missing* | 224 (3.1%) | 154 (5.4%) |
| **Education** |  |  |
| *Lower* | 2,605 (36.5%) | 427 (15%) |
| *Secondary* | 2,053 (28.8%) | 1,416 (49.8%) |
| *Higher* | 1,841 (25.8%) | 1,002 (35.2%) |
| *Other* | 634 (8.9%) | - |
| *Missing* | <10 (<0.5%) | <10 (<0.5%) |
| **Wealth quintiles** |  |  |
| *1 (Lowest)* | 1,095 (15.3%) | - |
| *2* | 1,298 (18.2%) | - |
| *3* | 1,439 (20.2%) | - |
| *4* | 1,537 (21.5%) | - |
| *5 (Highest)* | 1,659 (23.3%) | - |
| *Missing* | 108 (1.5%) | - |
| **Basic and instrumental activities of daily living** |  |  |
| *None* | 3,121 (43.7%) | 2,264 (79.5%) |
| *At least one* | 4,015 (56.3%) | 474 (16.7%) |
| *Missing* | - | 108 (3.8) |
| **Cardiovascular disease** |  |  |
| *No* | 6,340 (88.9%) | 2,092 (73.5%) |
| *Yes* | 795 (11.1%) | 754 (26.5%) |
| *Missing* | <10 (<0.5%) | - |
| **Depression symptoms at baseline (binary)** |  |  |
| *No depression* | 6,085 (85.3%) | 2,286 (80.3%) |
| *Depression* | 1,043 (14.6%) | 438 (15.4%) |
| *Missing* | <10 (<0.5%) | 122 (4.3%) |
| **Smoking status** |  |  |
| *Never smoked* | 2,636 (36.9%) | 1,309 (46%) |
| *Previous or current smoker* | 4,500 (63.1%) | 1,517 (53.3%) |
| *Missing* | - | 20 (0.70%) |
| **Alcohol intake** |  |  |
| *Monthly or less (ELSA)/no or occasional (SNAC-K)* | 2,101 (29.4%) | 958 (33.7%) |
| *Around weekly (ELSA)/light-to-moderate (SNAC-K)* | 2,276 (31.9%) | 1,410 (49.5%) |
| *Almost daily (ELSA)/heavy drinking (SNAC-K)* | 2,064 (28.9%) | 465 (16.3%) |
| *Not at all in last 12 months (ELSA)* | 694 (9.7%) | - |
| *Missing* | <10 (<0.5%) | 13 (0.5%) |
| **Physical activity** |  |  |
| *Vigorous activity at least once a week* | 2,198 (30.8%) | 617 (21.7%) |
| *Moderate activity at least once a week* | 3,479 (48.8%) | 1,391 (48.9%) |
| *Inactive (no moderate or vigorous activity)* | 1,456 (20.4%) | 838 (29.4%) |
| *Missing* | <10 (<0.5%) | - |
| **Social health markers** |  |  |
| **Marital/cohabitation status** |  |  |
| *Unmarried and alone* | 1,494 (20.9%) | 1,469 (51.6%) |
| *Married or cohabiting* | 5,642 (79.1%) | 1,373 (48.2%) |
| *Missing* | - | <10 (<0.5%) |
| **Contact frequency with family and friends** |  |  |
| *Never - every few months* | 3,347 (46.9%) | 341 (12.0%) |
| *Once or twice a month* | 2,555 (35.8%) | 1,186 (41.7%) |
| *One - three or more times per week* | 907 (12.7%) | 1,140 (40.1%) |
| *Missing* | 327 (4.6%) | 179 (6.3%) |
| **Social network size (continuous)** |  |  |
| *Range* | 0-30 | - |
| *Mean (SD)* | 7 (4.7) | - |
| *Missing* | 370 (5.2%) | - |
| **Social network size (categorical)** |  |  |
| *None* | 165 (2.3%) | 40 (1.4%) |
| *1-2 people* | 814 (11.4%) | 450 (15.8%) |
| *3-6 people* | 2,832 (39.7%) | 1,167 (41%) |

|  |  |  |  |
| --- | --- | --- | --- |
|  | 6 or more people | 2,955 (41.4%) | 910 (32%) |
|  | Missing | 370 (5.2%) | 279 (9.8%) |
| **Positive support (higher scores indicate more positive support)** |  |  |  |
|  | Range | 0-9 | 0-6 |
|  | Mean (SD) | 6.8 (1.6) | 5.3 (1.2) |
|  | Missing | 337 (4.7%) | 248 (8.7%) |
| **Negative support (higher scores indicate less negative support)** |  |  |  |
|  | Range | 0-9 | - |
|  | Mean (SD) | 7.1 (1.4) | - |
|  | Missing | 346 (4.9%) | - |
| **Cognition (baseline descriptives and rate of decline over time)<sup>1</sup>** |  |  |  |
| **Verbal fluency** |  |  |  |
|  | Range | 0-50 | 1-50 |
|  | Mean (SD) | 20.1 (6.2) | 20.5 (7.0) |
|  | Missing | <10 (<0.5%) | 23 (0.8%) |
| | Rate of decline, $\beta$ (95% CI)<sup>2</sup> | -0.016 (-0.018 - -0.013) | -0.053 (-0.061 - -0.045) |
| **Immediate recall** |  |  |  |
|  | Range | 0-10 | 0-16 |
|  | Mean (SD) | 5.7 (1.7) | 6.8 (2.5) |
|  | Missing | 23 (0.81%) | 74 (2.6%) |
| | Rate of decline, $\beta$ (95% CI)<sup>2</sup> | -0.028 (-0.031 - -0.026) | -0.054 (-0.063 - -0.044) |
| **Delayed recall** |  |  |  |
|  | Range | 0-10 | - |
|  | Mean (SD) | 4.3 (2.0) | - |
|  | Missing | <10 (<0.5%) | - |
| | Rate of decline, $\beta$ (95% CI)<sup>2</sup> | -0.036 (-0.038 - -0.034) | - |

<sup>1</sup>Raw cognitive scores presented for descriptives. Standardized outcomes used in main analysis.
<sup>2</sup>Rate of decline from linear mixed models.

**Table 2.**
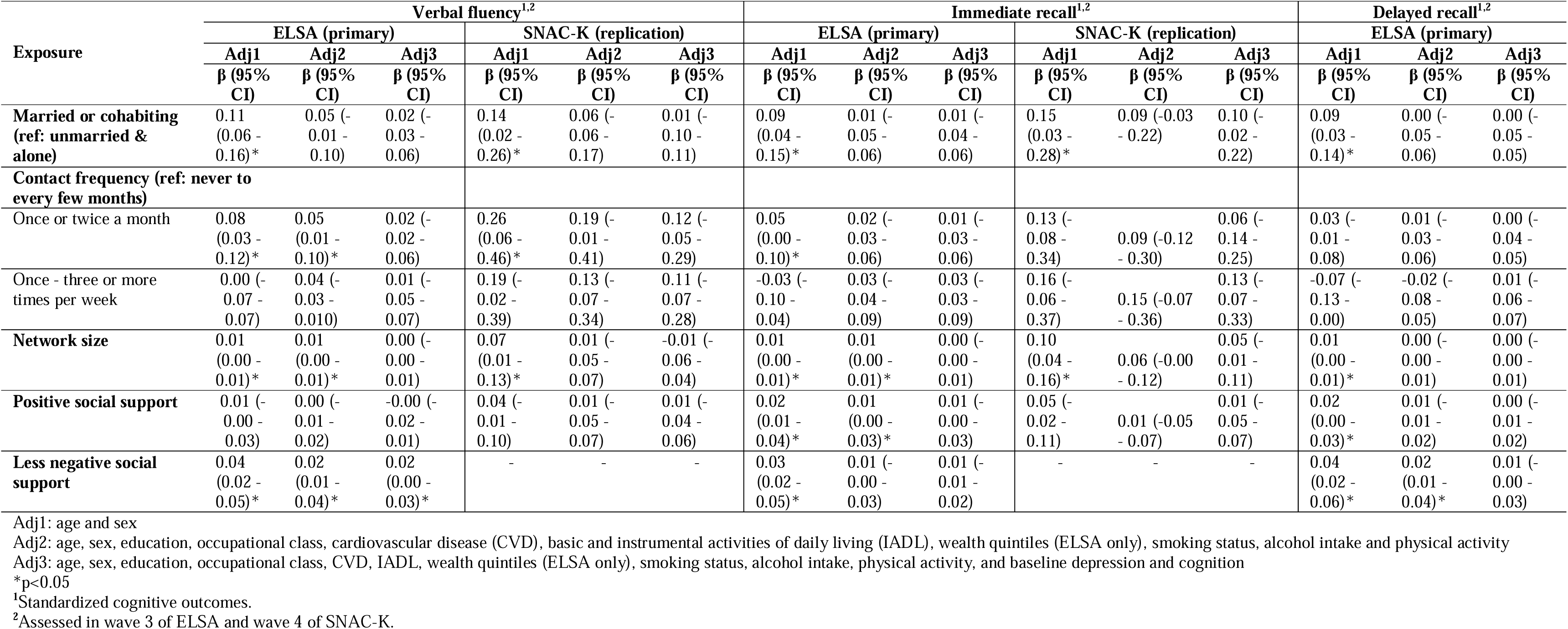
Associations between social health markers and subsequent cognition (single time point)

**Table 3.**
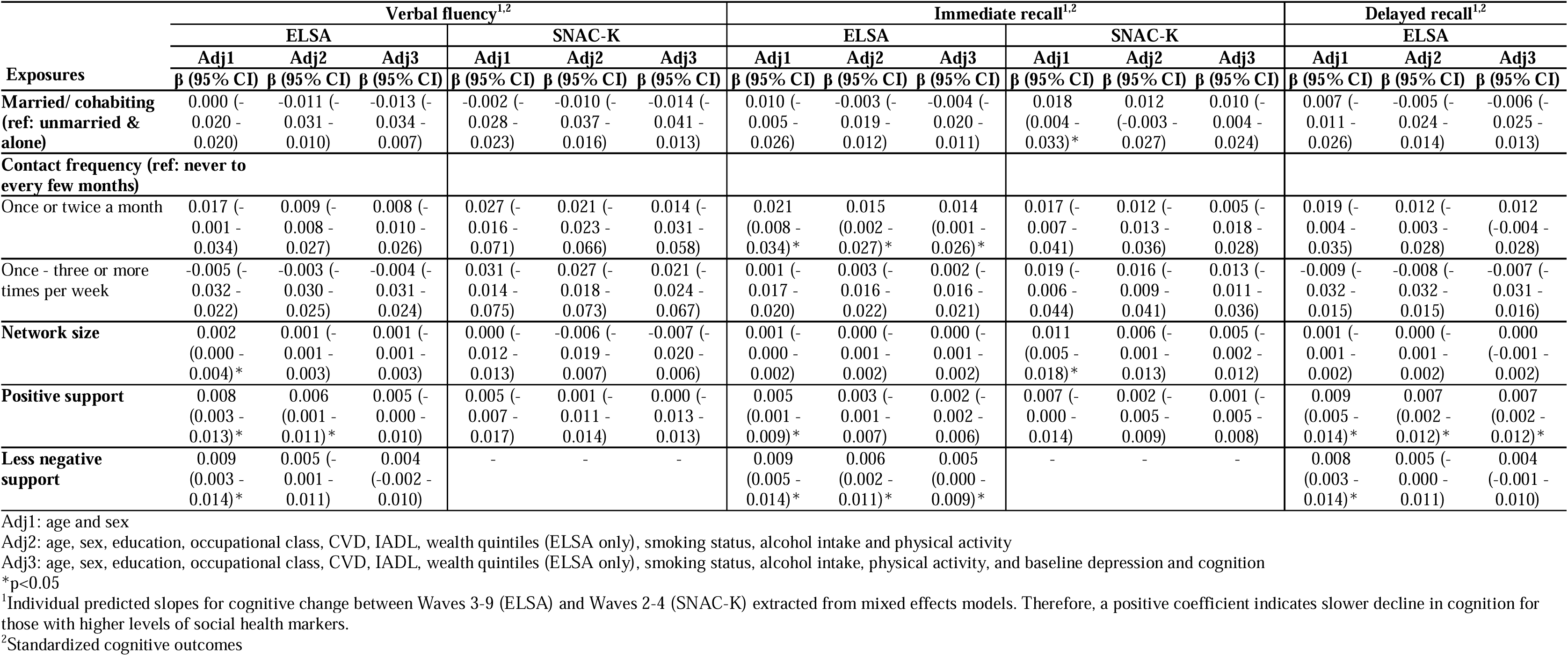
Associations between social health markers and subsequent cognitive change.

Depressive symptoms were tested as potential mediators, due to associations with social health markers after full adjustment, except contact frequency (Table S2), and with lower cognitive scores and faster decline across domains (Tables S3-S4). Inflammatory biomarkers were not tested as mediators due to limited association with exposures and outcomes.

#### Four-way decomposition - cognition (single time point)

In fully adjusted models, we found a total effect (TE) of network size on verbal fluency (0.01 (95%CI: 0.00 – 0.03) with an indirect effect via depressive symptoms (PIE=0.001, 95%CI: 0.000 – 0.001; Figure 1; Table S5). We found indirect effects of positive support on verbal fluency via depressive symptoms (PIE=0.002, 95%CI: 0.000 – 0.003), and on immediate recall (PIE=0.002, 95%CI: 0.001 – 0.003), where a TE was observed (0.01, 95%CI: 0.00 – 0.02). Negative support had a TE on verbal fluency (0.02, 95%CI: 0.00 – 0.03), and an indirect effect via depressive symptoms (PIE=0.002, 95%CI: 0.00 – 0.01). Mediation of associations between network size and negative support with recall outcomes, and of marital/cohabitation status on cognition, were not observed.

**Figure 1:**
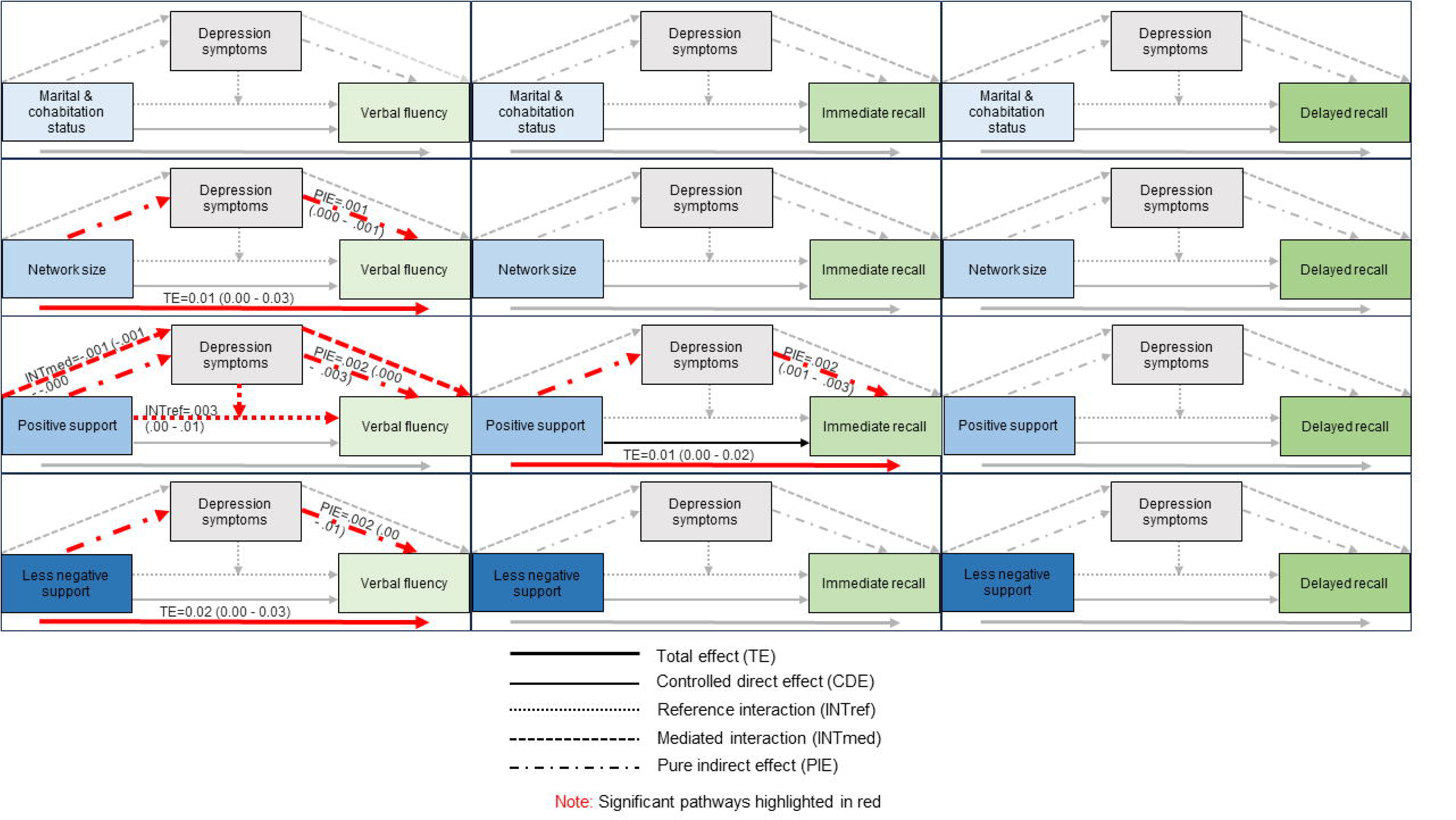
Four-way decomposition of total effects of social health on subsequent cognition (single time point) in ELSA - role of depressive symptoms.

#### Four-way decomposition - cognitive trajectories

We found an indirect effect of positive support on change in immediate recall via depressive symptoms (PIE= 0.001, 95%CI: 0.000 – 0.001; Figure 2; Table S6). Positive support had a TE on delayed recall (TE=0.005, 95%CI: 0.001 – 0.009) and a CDE (0.005, 95%CI: 0.001 – 0.009), with no evidence of mediation. Less negative support had a TE on immediate recall (0.006, 95%CI: 0.001 – 0.011), with an indirect effect via depressive symptoms (PIE=0.001, 95%CI: 0.000 – 0.002), which was also observed for delayed recall (PIE=0.001, 95%CI: 0.000 – 0.002). We found reference interactions indicating combined effects of network size and depressive symptoms on change in immediate (INTref=-0.001, 95%CI: −0.003 – −0.000) and delayed recall (INTref= −0.002, 95%CI: −0.003 – −0.000), although no TEs were found.

**Figure 2:**
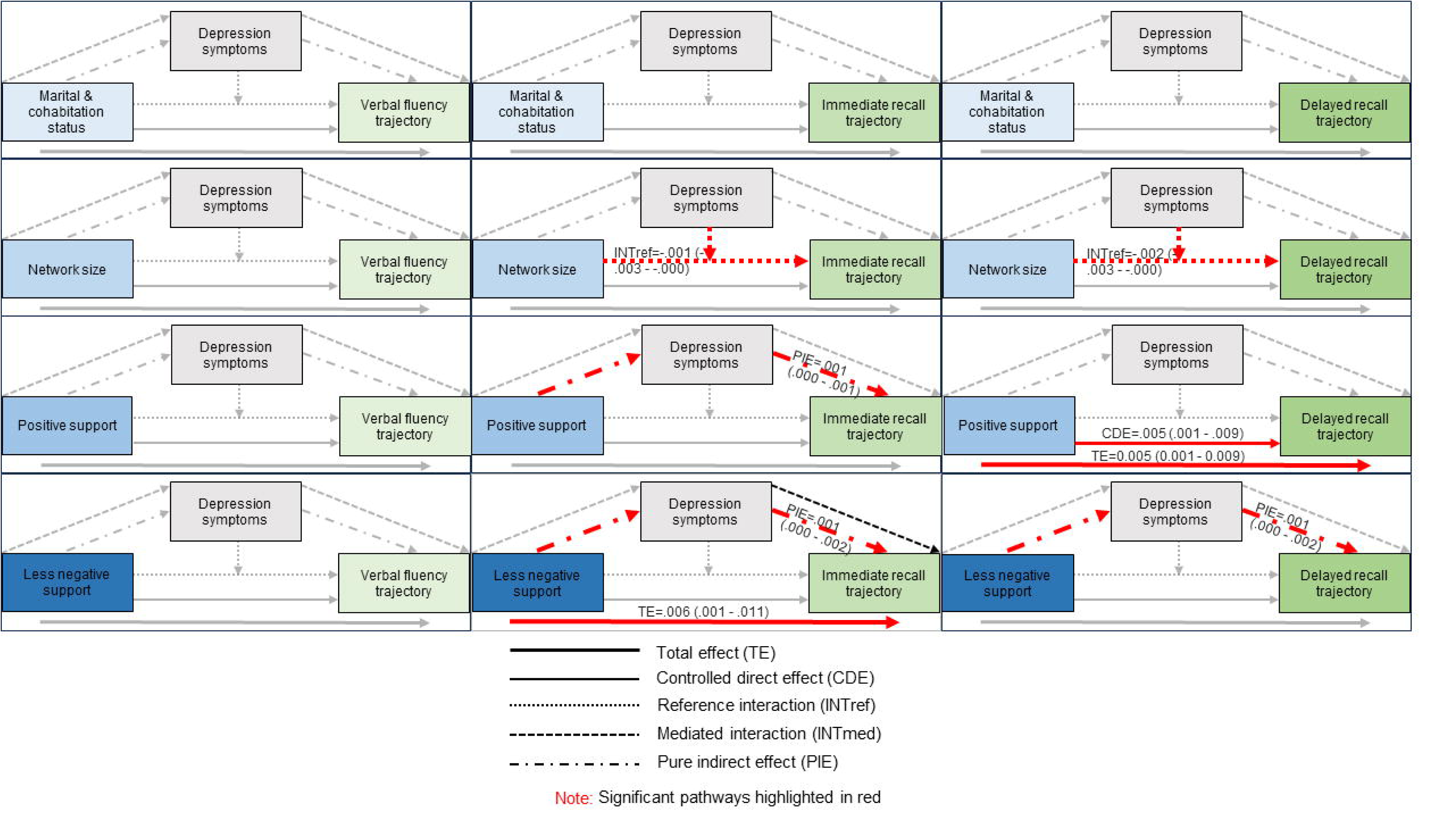
Four-way decomposition of total effects of social health on cognitive change in ELSA - role of depressive symptoms.

#### Sensitivity analyses

Social health remained associated with lower depression indexed as a binary variable (Table S7) and binary depression remained associated with lower cognitive scores and faster decline (Tables S3-4). Similar patterns of association were observed between CRP, exposures and outcomes when excluding those with CRP>10mg/L (Tables S3-S4, Table S8).

### Replication sample – SNAC-K

Descriptives for SNAC-K (n=2,846) are provided in Table 1 and Tables S1. Compared to ELSA, SNAC-K had an older mean baseline age (73.5, SD: 10.6 vs 63.8, SD: 9.4), a higher proportion of females (62.9% vs 55.5%) and a higher proportion of people with non-manual occupations (77.1% vs 59%). Associations between exposures, mediators and outcomes are reported in Tables 2-3 and Tables S2-4.

#### Four-way decomposition – cognition (single time point)

Indirect effects of positive support on verbal fluency via depressive symptoms found in ELSA were replicated in SNAC-K (PIE=0.003, 95%CI: 0.00 – 0.01; Table S5; Figure S4). In contrast, indirect effects of positive support on immediate recall, and network size on verbal fluency by depressive symptoms found in ELSA were not replicated in SNAC-K. In SNAC- K, there was an indirect effect of marital/cohabitation status on verbal fluency via depressive symptoms (PIE=0.02, 95%CI: 0.00 – 0.04) which was not found in ELSA. It was not possible to examine replication of associations relating to negative support or delayed recall.

#### Four-way decomposition – cognitive trajectories

The indirect effect of positive support on immediate recall change via depressive symptoms found in ELSA was replicated in SNAC-K (PIE=0.0003, 95%CI: 0.000 – 0.001; Table S6; Figure S5). We also found indirect effects of positive support on verbal fluency via depressive symptoms (PIE=0.001, 95%CI: 0.000 – 0.001; Table S6) and of marital/cohabitation status on verbal fluency change (PIE=0.005, 95%CI: 0.001 – 0.012) not found in ELSA.

## Discussion

### Summary and meaning of findings

Although positive social health markers have been posited to protect against cognitive decline, the pathways underpinning these associations are under-examined. We investigated the mediating roles of depressive symptoms and inflammation in associations between social health and subsequent cognition in ELSA, examining replication in SNAC-K. Using causal mediation analysis, we identified indirect effects via depressive symptoms of network size, positive and negative support on subsequent verbal fluency, and positive support on immediate recall. The positive support-verbal fluency mediation finding was replicated in SNAC-K, whereas the other relationships were not replicated or could not be tested. In SNAC-K, depressive symptoms mediated associations between marital/cohabitation status and verbal fluency, including trajectories, which was not observed in ELSA.

In ELSA, depressive symptoms also partially mediated effects of positive and negative support on immediate recall trajectories, and negative support on delayed recall trajectories. Although we observed total and controlled direct effects of positive support on delayed recall trajectories, we did not observe mediation, implying that this relationship did not operate via depressive symptoms. The positive support-immediate recall trajectory mediation finding was replicated in SNAC-K, whereas findings on delayed recall and negative support could not be tested in SNAC-K. Different relationships observed across cognitive domains and social health markers highlight the importance of examining associations individually.^34^

Our findings correspond with a previous study which found that depressive symptoms, but not anxiety, mediated the association between loneliness and subsequent cognitive functioning in the Irish Longitudinal Study on Ageing, although the indirect effect was small relative to the direct effect.^22^ Similarly, in the National Social Life, Health and Aging study, depression symptoms, among other factors such as functional ability, mediated associations between loneliness and general cognitive ability 10 years later.^20^

These results provide new insights into potential mechanisms linking social health with cognition, suggesting that interactional aspects of social health in particular, including more positive and less negative social support, may help to buffer against cognitive decline partly by lowering depressive symptoms. This supports the social bonding theory, which posits that closer social connections can impact health, including cognitive outcomes, by providing psychological benefits which then positively impact cognition.^35^

Establishing mechanisms linking social health and cognition is crucial for identifying possible targets for interventions to mitigate against cognitive decline, which is a major priority for public health. Despite the relatively modest effect sizes observed in this study, our findings could have significant implications at a population level, particularly given the high prevalence of depression and cognitive impairment in the population. Given that depressive symptoms did not fully mediate associations between social support and cognition, and the mixed findings regarding structural markers, further research is needed to examine other potential pathways through which social health could impact cognition.

Several key findings observed in ELSA were replicated in SNAC-K, particularly in relation to mediation of associations between positive support and cognition by depressive symptoms. However, findings differed between samples regarding mediation of associations between structural social health markers and cognition. It is possible that these discrepancies reflect differences across settings in relationships between social health, depression symptoms and cognition. For instance, factors such as marital and cohabitation status and network size could be interpreted and experienced differently across countries, thereby altering downstream relationships with subsequent depression and cognition. Discrepant findings could also reflect methodological differences between SNAC-K and ELSA, as described below.

We found no evidence of mediation by inflammatory biomarkers. These results align with two previous studies which did not find a mediating role of inflammatory biomarkers in associations between loneliness and global cognition.^20,25^An additional study involving older people in the United States found that CRP and fibrinogen partially mediated the association between social isolation and cognition in older men only.^26^ Further research is needed to investigate a wider range of inflammatory biomarkers and other possible physiological mechanisms, particularly those such as hypothalamic-pituitary-adrenal axis abnormalities, which may be influenced by the stress-buffering qualities of social relationships.^35^

### Strengths and limitations

Strengths include the use of data from two large, representative longitudinal cohorts, allowing examination of prospective associations and replication across samples. Few previous studies have investigated mediators of longitudinal associations between social health and cognition, despite longstanding hypotheses about mechanisms including mental health and physiological markers.

We applied causal mediation analysis, mitigating against some of the limitations of traditional mediation approaches, such as the assumed absence of exposure-mediator interaction. We investigated a range of structural and interactional social health exposures, and multiple cognitive domains. In contrast to most previous studies in this area, we examined mediation in relation to cognitive trajectories, in addition to cognition at a single time point. Studies included rich socio-demographic and health-related data, enabling adjustment for a range of potential confounders. Both studies had relatively long follow-up periods, helping to mitigate against reverse causality.

Nonetheless, we note several limitations. First, some measurements were not available in both samples. For instance, SNAC-K had a lower sensitivity measure of CRP than ELSA and did not include a measure of fibrinogen. In addition, SNAC-K did not have measures of delayed recall or negative social support. Although we sought to include comparable exposure, mediator and outcome measures across studies, it is possible that differences in results observed between ELSA and SNAC-K could reflect variation in measurement between studies. SNAC-K also had an older mean age at baseline than ELSA, and durations between exposures, mediators and outcomes differed between samples.

In addition, causal mediation analysis relies on assumptions about identification of and adjustment for exposure-mediator-outcome confounders. Although we adjusted for a range of important socio-demographic and health related confounders, we were not able to account for other potential confounders such as hearing loss and traumatic brain injury, and we cannot exclude the possibility of unmeasured confounders resulting in biased estimates. We also made several assumptions about temporal ordering of associations between social health markers with subsequent inflammation and depressive symptoms, as set out in our directed acyclic graphs. In addition, despite relatively long follow-up periods and adjustment for baseline depression and cognition, reverse causality remains a potential issue, whereby depressive symptoms, inflammation, and changes in social health could reflect early signs, rather than causes, of cognitive decline.

Although we investigated a range of social health and cognitive domains, further research is needed to examine these findings in relation to other outcomes, such as executive functioning, and other social health markers, including loneliness, which was not available at baseline in the present study. While examining replication was a strength, we were only able to compare results qualitatively, due to a lack of available methods to quantitatively assess replication between two studies. Further examination of replication is required across a wider range of settings, including in non-European cohorts.

### Conclusions

Our findings indicate that depressive symptoms are a pathway through which social health, particularly positive and negative aspects of social support, could influence subsequent cognitive outcomes, including cognitive decline. These insights into underlying mechanisms could contribute to the development of interventions and preventative strategies targeting social health, with potential downstream benefits for both mental health and cognitive functioning in older people.

## Data Availability

ELSA data used in this study are available to download through the UK Data Service. SNAC-K data used in this study are available to researchers upon approval by the SNAC-K data management and maintenance committee. Applications for accessing these data can be submitted to Maria Wahlberg at the Aging Research Centre, Karolinska Institutet, Stockholm, Sweden.

## Notes

### Competing Interest Statement

The authors have declared no competing interest.

### Funding Statement

This work was supported by the Social Health and Reserve in the Dementia patient journey (SHARED) SHARED Consortium, an EU Joint Programme Neurodegenerative Disease Research (JPND). The project is supported by Alzheimers Society (Ref:469) in the UK, by ZonMw/ JPND (733051082) in the Netherlands, by National Center for Research and Development in Poland, project number (NCBiR; JPND/06/2020) in Poland, by the National Health and Medical Research Council (NHMRC; APP1169489) in Australia. The English Longitudinal Study of Ageing was developed by a team of researchers based at University College London, NatCen Social Research, the Institute for Fiscal Studies, the University of Manchester, and the University of East Anglia. The data were collected by NatCen Social Research. The funding is currently provided by the National Institute on Aging (Ref: R01AG017644) and by a consortium of UK government departments: Department for Health and Social Care; Department for Transport; Department for Work and Pensions, which is coordinated by the National Institute for Health Research (NIHR, Ref: 198-1074). Funding has also been provided by the Economic and Social Research Council (ESRC). SNAC-K (http://www.snac.org) is financially supported by the Swedish Ministry of Health and Social Affairs; participating County Councils and Municipalities; the Swedish Research Council; and Swedish Research Council for Health, Working Life and Welfare. This project was funded by the Swedish Research Council for Health, Working Life, and Welfare (FORTE Grant No. 2018-01888 to AK Welmer).

## References

1 Vernooij-Dassen M, Verspoor E, Samtani S, et al. Conceptual advancement: Social health as a facilitator in the use of cognitive reserve. medRxiv 2022. DOI:10.1101/2022.06.07.22276079.

2 Santini ZI, Koyanagi A, Tyrovolas S, Mason C, Haro JM. The association between social relationships and depression: A systematic review. J Affect Disord 2015; 175: 53–65.

3 Valtorta NK, Kanaan M, Gilbody S, Ronzi S, Hanratty B. Loneliness and social isolation as risk factors for coronary heart disease and stroke: Systematic review and meta-analysis of longitudinal observational studies. Heart 2016; 102: 1009–16.

4 Holt-Lunstad J, Smith TB, Layton JB. Social relationships and mortality risk: A meta-analytic review. PLoS Med 2010; 7: e100316.

5 Kuiper JS, Zuidersma M, Oude Voshaar RC, et al. Social relationships and risk of dementia: A systematic review and meta-analysis of longitudinal cohort studies. Ageing Res Rev 2015; 22: 39–57.

6 Kelly ME, Duff H, Kelly S, et al. The impact of social activities, social networks, social support and social relationships on the cognitive functioning of healthy older adults: A systematic review. Syst Rev 2017; 6: 1–18.

7 Lenart-Bugla M, Łuc M, Pawłowski M, et al. What do we know about social and non-social factors influencing the pathway from cognitive health to dementia? A systematic review of reviews. Brain Sci. 2022; 12: 1214.

8 Samtani S, Mahalingam G, Lam BCP, et al. Associations between social connections and cognition: a global collaborative individual participant data meta-analysis. Lancet Healthy Longevity 2022; 3: e740–53.

9 Mahalingam G, Samtani S, Lam BCP, et al. Social connections and risk of incident mild cognitive impairment, dementia, and mortality in 13 longitudinal cohort studies of ageing. Alzheimer’s and Dementia 2023. DOI:10.1002/alz.13072.

10 Maddock J, Gallo F, Wolters FJ, et al. Social health and change in cognitive capability among older adults: findings from four European longitudinal studies. Gerontology 2023. DOI:10.1159/000531969.

11 Cohen S, Gottlieb BH, Underwood LG. Social relationships and health: challenges for measurement and intervention. Adv Mind Body Med 2001; 17: 129–41.

12 Santini ZI, Nielsen L, Hinrichsen C, et al. Social disconnectedness, perceived isolation, and symptoms of depression and anxiety among older Americans (NSHAP): a longitudinal mediation analysis. Articles Lancet Public Health 2020; 5: e62–70.

13 Cornwell EY, Waite LJ. Social Disconnectedness, Perceived Isolation, and Health among Older Adults. J Health Soc Behav 2009; 50: 31–48.

14 Santabárbara J, Lopez-Anton R, de la Camara C, et al. Clinically significant anxiety as a risk factor for dementia in the elderly community. Acta Psychiatr Scand 2019; 139: 6–14.

15 Gulpers B, Ramakers I, Hamel R, Köhler S, Voshaar RO, Verhey F. Anxiety as a Predictor for Cognitive Decline and Dementia: A Systematic Review and Meta-Analysis. The American Journal of Geriatric Psychiatry 2016; 24: 823–42.

16 Stafford J, Chung WT, Sommerlad A, Kirkbride JB, Howard R. Psychiatric disorders and risk of subsequent dementia: Systematic review and meta-analysis of longitudinal studies. Int J Geriatr Psychiatry. 2022; 37: 1–22.

17 Livingston G, Huntley J, Sommerlad A, et al. The Lancet Commissions Dementia prevention, intervention, and care: 2020 report of the Lancet Commission. The Lancet 2020; 396: 413–46.

18 Smith KJ, Gavey S, Riddell NE, Kontari P, Victor C. The association between loneliness, social isolation and inflammation: A systematic review and meta-analysis. Neurosci Biobehav Rev. 2020; 112: 519–41.

19 Peila R, Launer LJ. Inflammation and dementia: Epidemiologic evidence. Acta Neurol Scand 2006; 114: 102–6.

20 Kim AJ, Beam CR, Greenberg NE, Burke SL. Health Factors as Potential Mediators of the Longitudinal Effect of Loneliness on General Cognitive Ability. American Journal of Geriatric Psychiatry 2020; 28: 1272–83.

21 Peng C, Hayman LL, Mutchler JE, Burr JA. Friendship and Cognitive Functioning among Married and Widowed Chinese Older Adults. Journals of Gerontology - Series B Psychological Sciences and Social Sciences 2022; 77: 567–76.

22 McHugh Power J, Tang J, Kenny RA, Lawlor BA, Kee F. Mediating the relationship between loneliness and cognitive function: the role of depressive and anxiety symptoms. Aging Ment Health 2020; 24: 1071–8.

23 Wang H, Yang C, Yao Y. Familial factors, depression and cognitive decline: A longitudinal mediation analysis based on latent growth modeling (LGM). Int J Methods Psychiatr Res 2022; 31.

24 Peng C, Burr JA, Han SH. Cognitive function and cognitive decline among older rural Chinese adults: the roles of social support, pension benefits, and medical insurance. Aging Ment Health 2023; 27: 771–9.

25 Yu K, Siang Ng TK. Investigating biological pathways underpinning the longitudinal association between loneliness and cognitive impairment. J Gerontol A Biol Sci Med Sci 2023; 78: 1417–26.

26 Qi X, Ng TKS, Wu B. Sex differences in the mediating role of chronic inflammation on the association between social isolation and cognitive functioning among older adults in the United States. Psychoneuroendocrinology 2023; 149: 106023.

27 O’Connor M, Spry E, Patton G, et al. Better together: Advancing life course research through multi-cohort analytic approaches. Adv Life Course Res. 2022; 53: 100499.

28 Radloff LS. The CES-D Scale: A self-report depression scale for research in the general population. Appl Psychol Meas 1977; 1: 385–401.

29 Vanderweele TJ, Vansteelandt S. Conceptual issues concerning mediation, interventions and composition. Stat Interface 2009; 2: 457–68.

30 Rijnhart JJM, Lamp SJ, Valente MJ, MacKinnon DP, Twisk JWR, Heymans MW. Mediation analysis methods used in observational research: a scoping review and recommendations. BMC Med Res Methodol 2021; 21: 1–17.

31 Valeri L, VanderWeele TJ. Mediation analysis allowing for exposure–mediator interactions and causal interpretation: Theoretical assumptions andi With SAS and SPSS macro. Psychol Methods 2013; 18: 137–50.

32 O’Rourke HP, MacKinnon DPM. Reasons for testing mediation in the absence of an intervention effect: A research imperative in prevention and intervention research. J Stud Alcohol Drugs 2018; 79: 71–81.

33 Discacciati A, Bellavia A, Lee JJ, Mazumdar M, Valeri L. Med4way: A Stata command to investigate mediating and interactive mechanisms using the four-way effect decomposition. Int J Epidemiol 2019; 48: 15–20.

34 Gow AJ, Corley J, Starr JM, Deary IJ. Which social network or support factors are associated with cognitive abilities in old age? Gerontology 2013; 59: 454–63.

35 Perry BL, McConnell WR, Coleman ME, Roth AR, Peng S, Apostolova LG. Why the cognitive “fountain of youth” may be upstream: Pathways to dementia risk and resilience through social connectedness. Alzheimer’s and Dementia 2022; 18: 934–41.

