## Supplementary Material for "Social health and subsequent cognitive functioning in people aged 50 years and above: examining the mediating roles of depressive symptoms and inflammatory biomarkers"

### Table S1. Descriptive statistics for potential mediators

|  | **ELSA** | **SNAC-K** |
| --- | --- | --- |
| **Depression symptoms (binary)^1^** | **N (%)** | **N (%)** |
| *No depression* | 6,072 (85.2%) | 1,479 (90.5%) |
| *Depression* | 1,051 (14.8%) | 156 (9.5%) |
| **Depression symptoms (continuous)^1^** |  |  |
| *Mean (SD)* | 1.5 (1.9) | 1.76 (2.86) |
| *Range* | 0-8 | 0-22 |
| **CRP (mg/L)^1,2^** |  |  |
| *Range* | 0.2-210 | 1-237 |
| *Median (interquartile range (IQR))* | 2.0 (0.9 - 4.1) | 2.0 (1.0 - 4.0) |
| **Fibrinogen (g/L)^1,2^** |  |  |
| *Range* | 1-8.9 | - |
| *Median (IQR)* | 3.1 (2.7-3.6) | - |

**^1^**Mediators assessed at intermediate time point between exposures and outcomes. **^2^**Raw CRP values presented for descriptive purposes. Log transformed CRP used in main analysis.

| Table S2. Associations between social health markers and potential mediators | | | | | | | | | | | | | | | |
| --- | --- | --- | --- | --- | --- | --- | --- | --- | --- | --- | --- | --- | --- | --- | --- |
| **Exposures** | **Depressive symptoms^1^** | | | | | | **CRP^1,2^** | | | | | | **Fibrinogen^1^** | | |
|  | **ELSA** | | | **SNAC-K** | | | **ELSA** | | | **SNAC-K** | | |  | **ELSA** |  |
|  | **Adj1** | **Adj2** | **Adj3** | **Adj1** | **Adj2** | **Adj3** | **Adj1** | **Adj2** | **Adj3** | **Adj1** | **Adj2** | **Adj3** | **Adj1** | **Adj2** | **Adj3** |
|  | **β (95% CI)** | **β (95% CI)** | **β (95% CI)** | **β (95% CI)** | **β (95% CI)** | **β (95% CI)** | **β (95% CI)** | **β (95% CI)** | **β (95% CI)** | **β (95% CI)** | **β (95% CI)** | **β (95% CI)** | **β (95% CI)** | **β (95% CI)** | **β (95% CI)** |
| **Married or cohabiting (ref: unmarried & alone)** | -0.49 (-0.62 - -0.36)* | -0.31 (-0.43 - -0.18)* | -0.19 (-0.31 - -0.08)* | -0.74 (-1.02 - -0.46)* | -0.71 (-1.00 - -0.43)* | -0.43 (-0.69 - -0.16)* | -0.09 (-0.17 - -0.01) | 0.01 (-0.07 - 0.10) | 0.02 (-0.06 - 0.10) | -0.05 (-0.14 - 0.03) | -0.05 (-0.14 - 0.04) | -0.03 (-0.12 - 0.06) | -0.05 (-0.11 - 0.00) | -0.01 (-0.06 - 0.04) | 0.00 (-0.06 - 0.05) |
| **Contact frequency (ref: never to every few months)** | | | |  |  |  |  |  |  |  |  |  |  |  |  |
| Once or twice a month | -0.19 (-0.29 - -0.09)* | -0.16 (-0.25 - -0.06)* | -0.08 (-0.17 -0.01) | -0.04 (-0.52 - 0.44) | -0.04 (-0.52 - 0.45) | 0.12 (-0.31 - 0.56) | 0.04 (-0.02 - 0.11) | 0.05 (-0.01 - 0.12) | 0.07 (0.00 - 0.13)* | 0.02 (-0.13 - 0.17) | 0.08 (-0.07 - 0.23) | 0.10  (-0.05 - 0.24) | -0.05 (-0.09 - -0.01)* | -0.04 (-0.08 - 0.16) | -0.03 (-0.07 - 0.01) |
| Once - three or more times per week | -0.11 (-0.25 - 0.04) | -0.15 (-0.29 - -0.01)* | -0.11 (-0.24 - 0.02) | -0.30 (-0.79 - 0.19) | -0.21 (-0.70 - 0.28) | 0.09 (-0.35 - 0.53) | 0.13 (0.04 - 0.23)* | 0.12 (0.03 - 0.22)* | 0.13 (0.03 - 0.22)* | 0.10 (-0.05 - 0.25) | 0.15 (0.00 - 0.30) | 0.17  (0.02 - 0.32)* | 0.02 (-0.04 - 0.08) | 0.01 (-0.06 - 0.07) | 0.01 (-0.06 - 0.07) |
| **Network size** | -0.03 (-0.04 - -0.02)* | -0.03 (-0.03 - -0.02)* | -0.02 (-0.02 - -0.01)* | -0.28 (-0.42 - -0.14)* | -0.27 (-0.41 - -0.12)* | -0.08 (-0.21 - 0.05) | 0.00 (-0.01 - 0.01) | 0.00 (-0.01 - 0.01) | 0.00 (-0.01 - 0.01) | 0.00 (-0.04 - 0.05) | 0.02 (-0.03 - 0.06) | 0.03 (-0.02 - 0.07) | -0.01 (-0.01 - -0.00)* | -0.01 (-0.01 - -0.00)* | 0.00 (-0.01 - 0.00) |
| **Positive social support** | -0.22 (-0.26 - -0.19)* | -0.20 (-0.23 - -0.17)* | -0.13 (-0.16 - -0.11)* | -0.49 (-0.62 - -0.36)* | -0.50 (-0.63 - -0.37)* | -0.28 (-0.41 - -0.16)* | -0.02 (-0.04 - 0.01) | -0.01 (-0.03 - 0.01) | -0.01 (-0.03 - 0.01) | -0.03 (-0.08 - 0.01) | -0.03 (-0.07 - 0.02) | -0.02 (-0.06 - 0.03) | -0.01 (-0.02 - 0.00) | -0.01 (-0.02 - 0.01) | -0.01 (-0.02 - 0.01) |
| **Less negative social support** | -0.27 (-0.31 - -0.23)* | -0.23 (-0.27 - -0.19)* | -0.16 (-0.20 - -0.13)* | - | - | - | -0.03 (-0.05 - -0.01)* | -0.01 (-0.04 - 0.01) | -0.01 (-0.03 - 0.01) | - | - | - | -0.02 (-0.03 - -0.00)* | -0.01 (-0.03 - 0.00) | -0.01 (-0.02 - 0.01) |
| Adj1: age and sex Adj2: age, sex, education, occupational class, wealth quintiles (ELSA only), CVD, and IADL Adj3: age, sex, education, occupational class, wealth quintiles (ELSA only), CVD, IADL, smoking status, alcohol intake, physical activity, & baseline depression (for depression outcome only)  *p<0.05 ^1^Mediators assessed at intermediate time point between exposures and outcomes  ^2^Log transformed CRP | | | | | | | | | | | | | | | |

| Table S3. Associations between mediators and cognitive outcomes (single time point) | | | | | | | | | | | | | | | | | | |
| --- | --- | --- | --- | --- | --- | --- | --- | --- | --- | --- | --- | --- | --- | --- | --- | --- | --- | --- |
|  | | **Verbal fluency^1^** | | | | | | | | **Immediate recall^1^** | | | | | | **Delayed recall^1^** | | |
|  | | **ELSA** | | | | | **SNAC-K** | | | **ELSA** | | | **SNAC-K** | | | **ELSA** | | |
|  |  | **Adj1** | | **Adj2** | **Adj3** | | **Adj1** | **Adj2** | **Adj3** | **Adj1** | **Adj2** | **Adj3** | **Adj1** | **Adj2** | **Adj3** | **Adj1** | **Adj2** | **Adj3** |
|  |  | **β (95% CI)** | | **β (95% CI)** | **β (95% CI)** | | **β (95% CI)** | **β (95% CI)** | **β (95% CI)** | **β (95% CI)** | **β (95% CI)** | **β (95% CI)** | **β (95% CI)** | **β (95% CI)** | **β (95% CI)** | **β (95% CI)** | **β (95% CI)** | **β (95% CI)** |
| **Depression symptoms** | | -0.06 (-0.07 - -0.05)* | | -0.03 (-0.04 - -0.02)* | -0.01 (-0.02 - -0.00)* | | -0.03 (-0.06 - -0.01)* | -0.03 (-0.06 - -0.01)* | -0.03 (-0.05 - -0.01)* | -0.06 (-0.07 - -0.04)* | -0.02 (-0.04 - -0.01)* | -0.01 (-0.03 - -0.00)* | -0.03 (-0.05 - 0.00) | -0.01 (-0.04 - 0.01) | -0.01 (-0.04 - 0.02) | -0.05 (-0.06 - -0.04)* | -0.02 (-0.03 - -0.01)* | -0.01 (-0.02 - 0.00) |
| **Depression (binary)** | | -0.23 (-0.29 - -0.17)* | | -0.12 (-0.18 - -0.06)* | -0.05 (-0.10 - 0.01) | | -0.38 (-0.61 - -0.15)* | -0.34 (-0.57 - -0.11)* | -0.38 (-0.59 - -0.18)* | -0.21 (-0.27 - -0.15)* | -0.08 (-0.14 - -0.02)* | -0.04 (-0.10 - 0.02) | -0.38 (-0.64 - -0.12)* | -0.26 (-0.52 - 0.01) | -0.24 (-0.51 - 0.02) | -0.19 (-0.25 - -0.13)* | -0.07 (-0.13 - -0.01)* | 0.00 (-0.06 - 0.05) |
| **CRP^2^** | | -0.06 (-0.08 - -0.04)* | | -0.02 (-0.04 - 0.01) | -0.02 (-0.04 - 0.00) | | -0.07 (-0.14 - 0.00) | -0.05 (-0.12 - 0.02) | -0.04 (-0.1 - 0.02) | -0.05 (-0.08 - -0.03)* | -0.01 (-0.03 - 0.02) | 0.00 (-0.03 - 0.02) | -0.01 (-0.08 - 0.07) | 0.02 (-0.06 - 0.09) | 0.01 (-0.06 - 0.08) | -0.06 (-0.08 - -0.04)* | -0.01 (-0.04 - 0.01) | -0.01 (-0.03 - 0.01) |
| **CRP (sensitivity)^2,3^** | | -0.05 (-0.08 - -0.02)* | | 0.00 (-0.03 - 0.03) | 0.00 (-0.03 - 0.02) | | -0.09 (-0.18 - -0.01)* | -0.08 (-0.16 - 0.01) | -0.06 (-0.13 - 0.01) | -0.06 (-0.09 - -0.03)* | -0.01 (-0.03 - 0.02) | 0.00 (-0.03 - 0.02) | 0.04 (-0.06 - 0.13) | 0.03 (-0.06 - 0.13) | 0.02 (-0.06 - 0.11) | -0.08 (-0.11 - -0.05)* | -0.03 (-0.05 - 0.00) | -0.02 (-0.04 - 0.01) |
| **Fibrinogen** | | -0.05 (-0.08 - -0.01)* | | 0.01 (-0.02 - 0.05) | 0.00 (-0.03 - 0.03) | | - | - | - | -0.03 (-0.07 - 0.00) | 0.03 (-0.00 - 0.07) | 0.03 (-0.00 - 0.06) | - | - | - | -0.04 (-0.08 - -0.01)* | 0.01 (-0.02 - 0.05) | 0.02 (-0.01 - 0.05) |
| Adj1: age and sex  Adj2: age, sex, education, occupational class, wealth quintiles (ELSA only), CVD, IADL, smoking status, alcohol intake and physical activity Adj3: age, sex, education, occupational class, wealth quintiles (ELSA only), CVD, IADL, smoking status, alcohol intake, physical activity, baseline cognition and depression (for depression mediators only)  *p<0.05  ^1^Standardized cognitive outcomes. Assessed in Wave 3 in ELSA and Wave 4 in SNAC-K.  ^2^Log transformed CRP. ^3^Excluding those with CRP>10mg/L. | | | | | | | | | | | | | | | | | | |

| Table S4. Associations between mediators and cognitive change | | | | | | | | | | | | | | | |
| --- | --- | --- | --- | --- | --- | --- | --- | --- | --- | --- | --- | --- | --- | --- | --- |
|  | **Verbal fluency^1,2^** | | | | | | **Immediate recall^1,2^** | | | | | | **Delayed recall^1,2^** | | |
|  | **ELSA** | | | **SNAC-K** | | | **ELSA** | | | **SNAC-K** | | | **ELSA** | | |
|  | **Adj1** | **Adj2** | **Adj3** | **Adj1** | **Adj2** | **Adj3** | **Adj1** | **Adj2** | **Adj3** | **Adj1** | **Adj2** | **Adj3** | **Adj1** | **Adj2** | **Adj3** |
|  | **β (95% CI)** | **β (95% CI)** | **β (95% CI)** | **β (95% CI)** | **β (95% CI)** | **β (95% CI)** | **β (95% CI)** | **β (95% CI)** | **β (95% CI)** | **β (95% CI)** | **β (95% CI)** | **β (95% CI)** | **β (95% CI)** | **β (95% CI)** | **β (95% CI)** |
| **Depression symptoms^3^** | -0.011 (-0.015 - -0.007)* | -0.007 (-0.012 - -0.003)* | -0.006 (-0.010 - -0.001)* | -0.007 (-0.011 - -0.002)* | -0.007 (-0.012 - -0.002)* | -0.007 (-0.012 - -0.002)* | -0.011 (-0.014 - -0.008)* | -0.006 (-0.009 - -0.003)* | -0.004 (-0.007 - -0.001)* | -0.004 (-0.007 - -0.002)* | -0.004 (-0.006 - -0.001)* | -0.003 (-0.006 - -0.000)* | -0.012 (-0.015 - -0.008)* | -0.007 (-0.011 - -0.003)* | -0.005 (-0.010 - -0.001)* |
| **Depression (binary)^3^** | -0.053 (-0.075 - -0.031)* | -0.034 (-0.056 - -0.011)* | -0.026 (-0.050 - -0.002)* | -0.070 (-0.115 - -0.025)* | -0.075 (-0.122 - -0.028)* | -0.082 (-0.132 - -0.032)* | -0.051 (-0.068 - -0.034)* | -0.028 (-0.045 - -0.010)* | -0.020 (-0.038 - -0.002)* | -0.046 (-0.072 - -0.020)* | -0.043 (-0.069 - -0.016)* | -0.037 (-0.064 - -0.009)* | -0.065 (-0.086 - -0.044)* | -0.043 (-0.065 - -0.021)* | -0.038 (-0.061 - -0.015)* |
| **CRP^3,4^** | -0.005 (-0.013 - 0.004) | 0.003 (-0.006 - 0.012) | 0.003 (-0.006 - 0.012) | -0.002 (-0.017 - 0.013) | -0.002 (-0.018 - 0.013) | -0.001 (-0.016 - 0.014) | -0.010 (-0.016 - -0.004)* | -0.002 (-0.008 - 0.005) | -0.001 (-0.008 - 0.005) | 0.003 (-0.005 - 0.011) | 0.006 (-0.003 - 0.014) | 0.004 (-0.004 - 0.012) | -0.006 (-0.014 - 0.001) | 0.002 (-0.006 - 0.010) | 0.003 (-0.005 - 0.011) |
| **CRP (sensitivity)^3-5^** | -0.010 (-0.021 - 0.000) | -0.003 (-0.014 - 0.008) | -0.003 (-0.014 - 0.008) | -0.008 (-0.027 - 0.010) | -0.007 (-0.027 - 0.012) | -0.005 (-0.024 - 0.014) | -0.012 (-0.020 - -0.005)* | -0.003 (-0.011 - 0.005) | -0.003 (-0.010 - 0.005) | 0.007 (-0.004 - 0.017) | 0.007 (-0.004 - 0.017) | 0.005 (-0.005 - 0.015) | -0.007 (-0.016 - 0.002) | 0.001 (-0.008 - 0.011) | 0.003 (-0.007 - 0.013) |
| **Fibrinogen^3^** | -0.012 (-0.026 - 0.001) | -0.003 (-0.017 - 0.011) | -0.004 (-0.019 - 0.010) | - | - | - | -0.021 (-0.031 - -0.011)* | -0.010 (-0.020 - -0.000)* | -0.011 (-0.021 - -0.001)* | - | - | - | -0.021 (-0.033 - -0.008)* | -0.010 (-0.023 - 0.003) | -0.010 (-0.023 - 0.003) |
| Adj1: age and sex  Adj2: age, sex, education, occupational class, wealth quintiles (ELSA only), CVD, IADL, smoking status, alcohol intake and physical activity Adj3: age, sex, education, occupational class, wealth quintiles (ELSA only), CVD, IADL, smoking status, alcohol intake, physical activity, baseline cognition and depression (for depression mediators only)  *p<0.05  ^1^Individual predicted slopes for cognitive change between Waves 3-9 (ELSA) and Waves 2-4 (SNAC-K) extracted from mixed effects models. Therefore, a negative coefficient indicates faster decline in cognition for those with higher levels of depressive symptoms and inflammatory biomarkers.  ^2^Standardized cognitive outcomes.  ^3^Mediators assessed at intermediate time point between social health markers and cognitive outcomes. ^4^Log transformed CRP. ^5^Excluding those with CRP>10mg/L. | | | | | | | | | | | | | | | |

### Table S5. Four-way decomposition - total effects of social health on cognition (single time point) - role of depressive symptoms^1,2^

| **Verbal fluency** | | | | | | | | | | | | | | |
| --- | --- | --- | --- | --- | --- | --- | --- | --- | --- | --- | --- | --- | --- | --- |
|  | **Marital and cohabitation status** | | | | **Network size** | | | | **Positive support** | | | | **Negative support** | |
|  | **ELSA** | | **SNAC-K** | | **ELSA** | | **SNAC-K** | | **ELSA** | | **SNAC-K** | | **ELSA** | |
|  | **Adj1** | **Adj2** | **Adj1** | **Adj2** | **Adj1** | **Adj2** | **Adj1** | **Adj2** | **Adj1** | **Adj2** | **Adj1** | **Adj2** | **Adj1** | **Adj2** |
| **Total effect** | 0.11 (0.06 - 0.17)* | 0.02 (-0.03 - 0.07) | 0.13 (0.01 - 0.25)* | 0.00 (-0.10 - 0.11) | 0.02 (0.01 - 0.03)* | 0.01 (0.00 - 0.03)* | 0.04 (0.01 - 0.08)* | -0.01 (-0.04 -0.03) | 0.01 (-0.00 - 0.02) | -0.00 (-0.01 - 0.01) | 0.02 (-0.00 - 0.03) | 0.01 (-0.01 - 0.02) | 0.04 (0.02 - 0.06)* | 0.02 (0.00 - 0.03)* |
| **Controlled direct effect** | 0.09 (0.03 - 0.14)* | 0.01 (-0.04 -0.06) | 0.10 (-0.02 - 0.22) | -0.00 (-0.11 - 0.10) | 0.02 (0.00 - 0.03)* | 0.01 (-0.00 - 0.02) | 0.04 (-0.00 - 0.08) | -0.01 (-0.04 - 0.03) | -0.00 (-0.01 - 0.01) | -0.01 (-0.02 - 0.00) | 0.01 (-0.01 - 0.03) | 0.00 (-0.01 - 0.02) | 0.03 (0.01 - 0.05)* | 0.01 (-0.00 - 0.03) |
| **Reference interaction** | -0.00 (-0.01 - 0.01) | 0.01 (-0.01 - 0.02) | 0.00 (-0.01 - 0.01) | -0.01 (-0.02 - 0.01) | 0.00 (-0.00 - 0.00) | 0.00 (-0.00 - 0.01) | -0.00 (-0.01 - 0.01) | -0.00 (-0.01 - 0.01) | 0.001 (0.000 - 0.002)* | 0.003 (0.00 - 0.01)* | 0.00 (-0.00 - 0.01) | 0.00 (-0.00 - 0.00) | 0.00 (-0.00 - 0.00) | 0.00 (-0.00 - 0.01) |
| **Mediated interaction** | 0.00 (-0.01 - 0.01) | -0.00 (-0.01 - 0.00) | 0.01 (-0.03 - 0.04) | -0.01 (-0.03 - 0.01) | -0.00 (-0.00 - 0.00) | -0.00 (-0.00 -0.00) | -0.00 (-0.00 - 0.00) | -0.00 (-0.00 - 0.00) | -0.001 (-0.002 - -0.000)* | -0.001 (-0.001 - -0.000)* | 0.00 (-0.00 - 0.00) | 0.00 (-0.00 - 0.00) | -0.00 (-0.00 - 0.00) | -0.00 (-0.00 - 0.00) |
| **Pure indirect effect** | 0.03 (0.01 - 0.04)* | 0.00 (-0.00 - 0.01) | 0.02 (-0.00 - 0.05) | 0.02 (0.00 - 0.04)* | 0.004 (0.00 - 0.01)* | 0.001 (0.000 - 0.001)* | 0.00 (-0.00 - 0.01) | 0.00 (-0.00 - 0.01) | 0.01 (0.01 - 0.01)* | 0.002 (0.000 - 0.003)* | 0.003 (0.00 - 0.01)* | 0.003 (0.00 - 0.01)* | 0.02 (0.01 - 0.02)* | 0.002 (0.00 - 0.01)* |
| **Proportion mediated** | 0.24 (0.11 - 0.38)* | / | / | / | 0.21 (0.06 - 0.37)* | 0.04 (-0.02 - 0.09) | / | / | 1.19 (-0.54 - 2.92) | -1.05 (-10.13 - 8.03) | 0.24 (-0.07 - 0.55) | 0.54 (-1.01 - 2.09) | 0.35 (0.17 - 0.52)* | 0.11 (-0.05 - 0.26) |
| **Immediate recall** | | | | | | | | | | | | | | |
|  | **Marital and cohabitation status** | | | | **Network size** | | | | **Positive support** | | | | **Negative support** | |
|  | **ELSA** | | **SNAC-K** | | **ELSA** | | **SNAC-K** | | **ELSA** | | **SNAC-K** | | **ELSA** | |
|  | **Adj1** | **Adj2** | **Adj1** | **Adj2** | **Adj1** | **Adj2** | **Adj1** | **Adj2** | **Adj1** | **Adj2** | **Adj1** | **Adj2** | **Adj1** | **Adj2** |
| **Total effect** | 0.10 (0.04 - 0.15)* | 0.01 (-0.04 - 0.06) | 0.14 (0.02 - 0.27)* | 0.09 (-0.03 - 0.21) | 0.02 (0.00 - 0.03)* | 0.01 (-0.01 - 0.02) | 0.07 (0.03 - 0.11)* | 0.03 (-0.01 -0.07) | 0.02 (0.01 - 0.03)* | 0.01 (0.00 - 0.02)* | 0.02 (-0.00 - 0.03) | 0.00 (-0.01 - 0.02) | 0.04 (0.02 - 0.06)* | 0.01 (-0.01 - 0.02) |
| **Controlled direct effect** | 0.07 (0.02 - 0.13)* | 0.01 (-0.05 - 0.06) | 0.12 (-0.01 - 0.25) | 0.08 (-0.05 - 0.20) | 0.01 (-0.00 - 0.02) | 0.01 (-0.00 - 0.02) | 0.06 (0.02 - 0.10)* | 0.03 (-0.01 -0.07) | 0.01 (-0.00 - 0.02) | 0.01 (-0.00 - 0.02) | 0.01 (-0.01 - 0.03) | -0.00 (-0.02 - 0.02) | 0.02 (0.00 - 0.04)* | 0.01 (-0.01 - 0.02) |
| **Reference interaction** | -0.01 (-0.01 - 0.00) | 0.00 (-0.01 - 0.02) | 0.00 (-0.01 - 0.02) | 0.01 (-0.01 - 0.02) | -0.00 (-0.00 - 0.00) | -0.00 (-0.01 - 0.00) | 0.01 (-0.01 - 0.02) | 0.00 (-0.01 - 0.01) | 0.00 (-0.00 - 0.00) | 0.00 (-0.00 - 0.01) | 0.00 (-0.00 - 0.01) | 0.00 (-0.00 - 0.01) | -0.00 (-0.00 - 0.00) | -0.00 (-0.01 - 0.00) |
| **Mediated interaction** | 0.01 (-0.01 - 0.02) | -0.00 (-0.00 - 0.00) | 0.01 (-0.03 - 0.04) | 0.01 (-0.01 - 0.03) | 0.00 (-0.00 - 0.00) | 0.00 (-0.00 - 0.00) | 0.00 (-0.00 - 0.00) | 0.00 (-0.00 - 0.00) | -0.001 (-0.002 - -0.000)* | -0.00 (-0.00 - 0.00) | 0.00 (-0.00 - 0.00) | 0.00 (-0.00 - 0.00) | 0.00 (-0.00 - 0.00) | 0.00 (-0.00 - 0.00) |
| **Pure indirect effect** | 0.02 (0.01 - 0.03)* | 0.00 (-0.00 - 0.01) | 0.01 (-0.01 - 0.03) | -0.00 (-0.02 - 0.01) | 0.004 (0.00 - 0.01)* | 0.00 (-0.00 - 0.00) | 0.00 (-0.00 - 0.01) | -0.00 (-0.00 - 0.00) | 0.01 (0.01 - 0.01)* | 0.002 (0.001 - 0.003)* | 0.00 (-0.00 - 0.01) | 0.00 (-0.00 - 0.00) | 0.01 (0.01 - 0.02)* | 0.00 (-0.00 - 0.01) |
| **Proportion mediated** | 0.29 (0.10 - 0.47)* | / | / | / | 0.28 (0.03 - 0.53)* | / | / | / | 0.52 (0.16 - 0.88)* | 0.13 (-0.04 - 0.30) | / | / | 0.41 (0.18 - 0.64)* | / |
| **Delayed recall** | | | | | | | | | | | | | | |
|  | **Marital/cohabitation status** | | | | **Network size** | | | | **Positive support** | | | | **Negative support** | |
|  | **ELSA** | | **SNAC-K** | | **ELSA** | | **SNAC-K** | | **ELSA** | | **SNAC-K** | | **ELSA** | |
|  | **Adj1** | **Adj2** | **Adj1** | **Adj2** | **Adj1** | **Adj2** | **Adj1** | **Adj2** | **Adj1** | **Adj2** | **Adj1** | **Adj2** | **Adj1** | **Adj2** |
| **Total effect** | 0.09 (0.04 - 0.15)* | -0.01 (-0.06 - 0.04) | - | - | 0.01 (0.00 - 0.03)* | 0.01 (-0.01 - 0.02) | - | - | 0.01 (0.00 - 0.02)* | 0.00 (-0.01 - 0.01) | - | - | 0.05 (0.03 - 0.06)* | 0.01 (-0.01 - 0.03) |
| **Controlled direct effect** | 0.07 (0.02 - 0.13)* | 0.00 (-0.05 - 0.05) | - | - | 0.01 (-0.00 - 0.02) | 0.01 (-0.01 - 0.02) | - | - | 0.00 (-0.01 - 0.02) | 0.00 (-0.01 - 0.01) | - | - | 0.03 (0.01 - 0.05)* | 0.01 (-0.00 - 0.03) |
| **Reference interaction** | -0.01 (-0.02 - 0.00) | -0.01 (-0.02 - 0.01) | - | - | -0.00 (-0.00 - 0.00) | -0.00 (-0.01 - 0.00) | - | - | 0.00 (-0.00 - 0.00) | 0.00 (-0.00 - 0.00) | - | - | -0.00 (-0.00 - 0.00) | -0.00 (-0.01 - 0.00) |
| **Mediated interaction** | 0.01 (-0.00 - 0.02) | 0.00 (-0.00 - 0.01) | - | - | 0.00 (-0.00 - 0.00) | 0.00 (-0.00 - 0.00) | - | - | -0.00 (-0.00 - 0.00) | -0.00 (-0.00 - 0.00) | - | - | 0.00 (-0.00 - 0.00) | 0.00 (-0.00 - 0.00) |
| **Pure indirect effect** | 0.02 (0.01 - 0.03)* | -0.00 (-0.00 - 0.00) | - | - | 0.004 (0.00 - 0.01)* | 0.00 (-0.00 - 0.00) | - | - | 0.01 (0.01 - 0.01)* | 0.00 (-0.00 - 0.00) | - | - | 0.01 (0.01 - 0.02)* | 0.00 (-0.00 - 0.00) |
| **Proportion mediated** | 0.30 (0.10 - 0.49)* | / | - | - | 0.29 (0.01 - 0.56)* | / | - | - | 0.68 (0.09 - 1.26)* | / | - | - | 0.32 (0.17 - 0.48)* | / |
| Adj1: age and sex Adj2: age, sex, education, occupational class, wealth quintiles (ELSA only), CVD, IADL, smoking status, alcohol intake, physical activity, baseline cognition and depression  ^1^Cognitive outcomes assessed in Wave 3 in ELSA and Wave 4 in SNAC-K.  ^2^For marital and cohabitation status, people who were married and/or cohabiting were compared with those who were unmarried and living alone. For network size, positive and negative support, comparisons were made between the 25^th^ percentile and the mean value. | | | | | | | | | | | | | | |

| Table S6. **Four-way decomposition - total effects of social health on cognitive change - role of depressive symptoms^1,2^** | | | | | | | | | | | | | | |
| --- | --- | --- | --- | --- | --- | --- | --- | --- | --- | --- | --- | --- | --- | --- |
| **Social health exposure^1^** | | | | | | | | | | | | | | |
|  | **Verbal fluency trajectories** | | | | | | | | | | | | | |
|  | **Marital and cohabitation status** | | | | **Network size** | | | | **Positive support** | | | | **Negative support** | |
|  | **ELSA** | | **SNAC-K** | | **ELSA** | | **SNAC-K** | | **ELSA** | | **SNAC-K** | | **ELSA** | |
|  | **Adj1** | **Adj2** | **Adj1** | **Adj2** | **Adj1** | **Adj2** | **Adj1** | **Adj2** | **Adj1** | **Adj2** | **Adj1** | **Adj2** | **Adj1** | **Adj2** |
| **Total effect** | .001 (-.020 - .022) | -.015 (-.037 - .007) | -.004 (-.030 - .022) | -.017 (-.044 - .010) | .005 (-.000 - .010) | .001 (-.005 - .006) | .001 (-.008 - .009) | -.004 (-.013 - .004) | .007 (.003 - .011)* | .003 (-.001 - .008) | .002 (-.001 - .005) | .000 (-.003 - .004) | .009 (.003 - .016)* | .004 (-.003 - .011) |
| **Controlled direct effect** | -.003 (-.024 - .018) | -.013 (-.035 - .009) | -.010 (-.036 - .017) | -.019 (-.046 - .008) | .004 (-.001 - .009) | .001 (-.004 - .006) | -.000 (-.008 - .008) | -.004 (-.013 - .004) | .005 (.000 - .009)* | .004 (-.001 - .008) | .001 (-.003 - .004) | -.000 (-.004 - .003) | .006 (-.001 - .013) | .003 (-.004 - .010) |
| **Reference interaction** | -.002 (-.004 - .001) | -.002 (-.008 - .003) | .000 (-.002 - .002) | -.000 (-.002 - .002) | .000 (-.000 - .000) | -.001 (-.003 - .001) | -.000 (-.001 - .001) | -.001 (-.002 - .001) | -.000 (-.000 - .000) | -.001 (-.003 - .000) | .000 (-.000 - .001) | .000 (-.000 - .001) | -.000 (-.001 - .001) | -.000 (-.003 - .002) |
| **Mediated interaction** | .003 (-.002 - .007) | .001 (-.001 - .002) | -.001 (-.009 - .007) | -.002 (-.009 - .004) | .000 (-.000 - .000) | .000 (-.000 - .000) | -.000 (-.001 - .000) | -.000 (-.000 - .000) | .0004 (.000 - .001)* | .0003 (.000 - .001)* | .000 (-.000 - .000) | .000 (-.000 - .000) | .000 (-.001 - .001) | .000 (-.001 - .001) |
| **Pure indirect effect** | .003 (-.001 - .007) | .000 (-.001 - .002) | .006 (.001 - .012)* | .005 (.001 - .010)* | .001 (.000 - .001)* | .000 (-.000 - .000) | .001 (.000 - .003)* | .001 (-.000 - .002) | .002 (.001 - .003)* | .001 (-.000 - .001) | .001 (.000 - .002)* | .001 (.000 - .001)* | .003 (.002 - .005)* | .001 (-.000 - .002) |
| **Proportion mediated** | / | / | -1.43 (-11.22 - 8.36) | -0.17 (-0.60 - 0.27) | 0.18 (-0.03 – 0.38) | / | 1.74 (-21.07 - 24.55) | / | 0.35 (0.09 - 0.60)* | / | 0.46 (-0.34 - 1.25) | 1.65 (-12.19 - 15.49) | 0.37 (0.06 - 0.68)* | / |
|  | **Immediate recall trajectories** | | | | | | | | | | | | | |
|  | **Marital and cohabitation status** | | | | **Network size** | | | | **Positive support** | | | | **Negative support** | |
|  | **ELSA** | | **SNAC-K** | | **ELSA** | | **SNAC-K** | | **ELSA** | | **SNAC-K** | | **ELSA** | |
|  | **Adj1** | **Adj2** | **Adj1** | **Adj2** | **Adj1** | **Adj2** | **Adj1** | **Adj2** | **Adj1** | **Adj2** | **Adj1** | **Adj2** | **Adj1** | **Adj2** |
| **Total effect** | .011 (-.004 - .026) | -.006 (-.021 - .009) | .018 (.003 - .032* | .010 (-.004 - .024) | .002 (-.002 - .005) | -.001 (-.005 - .003) | .008 (.003 - .012)* | .004 (-.001 - .008) | .004 (.001 - .007)* | .001 (-.002 - .005) | .002 (.000 - .004)* | .001 (-.001 - .002) | .011 (.006 - .016)* | .006 (.001 - .011)* |
| **Controlled direct effect** | .007 (-.008 - .022) | -.005 (-.020 - .011) | .014 (-.001 - .028) | .007 (-.007 - .022) | .001 (-.003 - .004) | .000 (-.003 - .004) | .007 (.003 - .012)* | .003 (-.001 - .008) | .002 (-.001 - .005) | .001 (-.002 - .004) | .001 (-.001 - .003) | .001 (-.002 - .002) | .007 (.002 - .012)* | .005 (-.000 - .010) |
| **Reference interaction** | -.001 (-.003 - .001) | -.002 (-.006 - .002) | -.000 (-.001 - .001) | .000 (-.001 - .001) | .000 (-.000 - .000) | -.001 (-.003 - -.000)* | .000 (-.000 - .001) | .000 (-.001 - .001) | -.000 (-.000 - .000) | -.000 (-.001 - .001) | .000 (-.000 - .000) | .000 (-.000 - .000) | .000 (-.000 - .001) | .000 (-.001 - .002) |
| **Mediated interaction** | .002 (-.001 - .005) | .000 (-.001 - .002) | .001 (-.003 - .006) | .001 (-.002 - .004) | .000 (.000 - .000) | .000 (-.000 - .000) | .000 (-.000 - .000) | .000 (-.000 - .000) | .000 (-.000 - .000) | .000 (-.000 - .000) | .000 (-.000 - .000) | .000 (.000 - .000) | -.000 (-.001 - .001) | -.000 (-.001 - .000) |
| **Pure indirect effect** | .003 (.001 - .006)* | .000 (-.001 - .001) | .003 (-.001 - .006) | .001 (-.001 - .003) | .001 (.000 - .001)* | .000 (-.000 -.000) | .001 (-.000 - .001) | .000 (-.000 - .001) | .002 (.001 - .003)* | .001 (.000 - .001)* | .001 (.000 - .001)* | .0003 (.000 - .001)* | .003 (.002 - .005)* | .001 (.000 - .002)* |
| **Proportion mediated** | 0.49 (-0.20 – 1.18) | / | / | / | 0.46 (-0.46 – 1.38) | / | / | / | 0.51 (0.10 - 0.91)* | 0.38 (-0.52 – 1.29) | 0.28 (0.03 - 0.58)* | 0.55 (-1.36 - 2.46) | 0.30 (0.13 - 0.47)* | 0.13 (-0.04 - 0.29)* |
|  | **Delayed recall trajectories** | | | | | | | | | | | | | |
|  | **Marital and cohabitation status** | | | | **Network size** | | | | **Positive support** | | | | **Negative support** | |
|  | **ELSA** | | **SNAC-K** | | **ELSA** | | **SNAC-K** | | **ELSA** | | **SNAC-K** | | **ELSA** | |
|  | **Adj1** | **Adj2** | **Adj1** | **Adj2** | **Adj1** | **Adj2** | **Adj1** | **Adj2** | **Adj1** | **Adj2** | **Adj1** | **Adj2** | **Adj1** | **Adj2** |
| **Total effect** | .007 (-.011 - .026) | -.007 (-.026 - .012) | - | - | .002 (-.003 - .006) | -.002 (-.006 - .003) | - | - | .008 (.004 - .012)* | .005 (.001 - .009)* | - | - | .009 (.003 - .015)* | .006 (-.001 - .012) |
| **Controlled direct effect** | .003 (-.016 - .022) | -.007 (-.026 - .013) | - | - | .001 (-.004 - .005) | -.000 (-.004 - .004) | - | - | .006 (.002 - .010)* | .005 (.001 - .009)* | - | - | .005 (-.001 - .012) | .004 (-.003 - .010) |
| **Reference interaction** | -.001 (-.003 - .002) | -.001 (-.006 - .004) | - | - | .000 (-.000 - .000) | -.002 (-.003 - -.000)* | - | - | -.000 (-.000 - .000) | -.001 (-.002 - .001) | - | - | .000 (-.000 - .001) | .001 (-.001 - .003) |
| **Mediated interaction** | .002 (-.002 - .005) | .000 (-.001 - .002) | - | - | .000 (-.000 - .000) | .000 (-.000 - .000) | - | - | .000 (-.000 - .001) | .000 (-.000 - .000) | - | - | -.001 (-.002 - .000) | -.000 (-.001 - .003) |
| **Pure indirect effect** | .004 (-.001 - .008) | .001 (-.001 - .002) | - | - | .001 (.000 - .001)* | .000 (-.000 - .000) | - | - | .002 (.001 - .003)* | .000 (-.000 - .001) | - | - | .004 (.002 - .005)* | .001 (.000 - .002)* |
| **Proportion mediated** | 0.71 (-1.00 – 2.42) | / | - | - | 0.51 (-0.87 – 1.89) | / | - | - | 0.28 (0.10 - 0.45)* | / | - | - | 0.36 (0.08 - 0.64)* | 0.14 (-0.08 - 0.37) |
| Adj1: age and sex Adj2: age, sex, education, occupational class, wealth quintiles (ELSA only), CVD, IADL, smoking status, alcohol intake, physical activity, baseline cognition and depression  ^1^Individual predicted slopes for cognitive change between Waves 3-9 (ELSA) and Waves 2-4 (SNAC-K) extracted from mixed effects models.  ^2^For marital and cohabitation status, people who were married and/or cohabiting were compared with those who were unmarried and living alone. For network size, positive and negative support, comparisons were made between the 25^th^ percentile and the mean value. | | | | | | | | | | | | | | |

### Table S7. Sensitivity analysis - associations between social health exposures and binary depression mediator

|  | **ELSA** **Odds ratio (OR)** | | | **SNAC-K**  **OR** | | |
| --- | --- | --- | --- | --- | --- | --- |
| **Exposures** | **Adj1** | **Adj2** | **Adj3** | **Adj1** | **Adj2** | **Adj3** |
| **Married or cohabiting (ref: unmarried & alone)** | 0.60 (0.51 - 0.70)* | 0.71 (0.60 - 0.85)* | 0.80 (0.66 - 0.96)* | 0.46 (0.32 - 0.67)* | 0.48 (0.33 - 0.71)* | 0.64 (0.42 - 0.96)* |
| **Contact frequency (ref: never to every few months)** | |  |  |  |  |  |
| Once or twice a month | 0.76 (0.65 - 0.88)* | 0.77 (0.66 - 0.91)* | 0.86 (0.72 - 1.01) | 0.78 (0.44 - 1.39) | 0.86 (0.47 - 1.59) | 1.16 (0.57 - 2.39) |
| Once - three or more times per week | 0.86 (0.70 - 1.06) | 0.80 (0.64 - 0.99)* | 0.83 (0.65 - 1.06) | 0.58 (0.32 - 1.06) | 0.71 (0.38 - 1.34) | 1.09 (0.52 - 2.28) |
| **Network size** | 0.96 (0.95 - 0.98)* | 0.97 (0.95 - 0.99)* | 0.98 (0.97 - 1.00)* | 0.63 (0.53 - 0.76)* | 0.65 (0.52 - 0.79)* | 0.76 (0.60 - 0.95)* |
| **Positive support^1^** | 0.78 (0.75 - 0.82)* | 0.80 (0.77 - 0.84)* | 0.86 (0.82 - 0.90)* | 0.65 (0.56 - 0.74)* | 0.64 (0.56 - 0.74)* | 0.76 (0.64 - 0.89)* |
| **Negative support^1^** | 0.76 (0.72 - 0.79)* | 0.78 (0.74 - 0.82)* | 0.83 (0.79 - 0.88)* | / | / | / |
| Adj1: age and sex Adj2: age, sex, education, occupational class, wealth quintiles (ELSA only), CVD, and IADL Adj3: age, sex, education, occupational class, wealth quintiles (ELSA only), CVD, IADL, smoking status, alcohol intake, physical activity, & baseline depression (for depression outcome only) | | | | | | |

| Table S8. Sensitivity analysis - associations between social health exposures and CRP excluding those with CRP>10mg/L^1^ | | | | | | |
| --- | --- | --- | --- | --- | --- | --- |
| **Exposures** | **ELSA** | | | **SNAC-K** | | |
|  | **Adj1** | **Adj2** | **Adj3** | **Adj1** | **Adj2** | **Adj3** |
|  | **β (95% CI)** | **β (95% CI)** | **β (95% CI)** | **β (95% CI)** | **β (95% CI)** | **β (95% CI)** |
| **Married/cohabiting (ref: unmarried & alone)** | -0.05 (-0.12 - 0.02) | 0.03 (-0.04 - 0.10) | 0.04 (-0.03 - 0.11) | -0.03 (-0.10 - 0.04) | -0.03 (-0.10 - 0.05) | -0.02 (-0.10 - 0.05) |
| **Contact frequency (ref: never to every few months)** | | | | | | |
| Once or twice a month | 0.05 (-0.01 - 0.11) | 0.07 (0.01 - 0.13)* | 0.08 (0.02 - 0.13)* | 0.01 (-0.11 - 0.13) | 0.04 (-0.08 - 0.16) | 0.05 (-0.07 - 0.17) |
| Once - three or more times per week | 0.07 (-0.01 - 0.15) | 0.06 (-0.03 - 0.14) | 0.06 (-0.02 - 0.15) | 0.06 (-0.07 - 0.18) | 0.08 (-0.05 - 0.20) | 0.08 (-0.04 - 0.21) |
| **Network size** | 0.00 (-0.01 -0.01) | 0.00 (-0.00 - 0.01) | 0.00 (-0.00 - 0.01) | -0.00 (-0.04 - 0.04) | 0.00 (-0.03 - 0.04) | 0.01 (-0.03 - 0.05) |
| **Positive social support^2^** | -0.01 (-0.03 - 0.01) | -0.00 (-0.02 - 0.02) | -0.00 (-0.02 - 0.02) | -0.04 (-0.07 - -0.00)* | -0.03 (-0.07 - 0.00) | -0.03 (-0.07 - 0.01) |
| **Less negative social support^2^** | -0.03 (-0.05 - -0.01)* | -0.02 (-0.04 - 0.01) | -0.01 (-0.03 - 0.01) | - | - | - |
| Adj1: age and sex Adj2: age, sex, education, occupational class, wealth quintiles (ELSA only), CVD, and IADL Adj3: age, sex, education, occupational class, wealth quintiles (ELSA only), CVD, IADL, smoking status, alcohol intake, and physical activity  *p<0.05 ^1^Log transformed. | | | | | | |

### Figure S1. Flow diagram - ELSA analytic sample

11,391 core participants aged over 50 years took part at baseline

353 participants excluded due to missing baseline data on:
- Cognition (no measures available) (n=353)
- All baseline social health variables (n=0)

100 participants excluded due to having recorded dementia at baseline or intermediate time point

11,038 participants with baseline data on cognitive function and social health

2,459 participants excluded for not having data on all potential mediators

10,938 participants without recorded dementia before wave 3

8,479 participants had data on at least one mediator in wave 2

1,343 participants excluded for not having data on any cognitive measure at wave 3 and the same measure in at least two subsequent time points

Analytic sample n=7,136

### Figure S2. Directed acyclic graph - depression symptoms as mediator

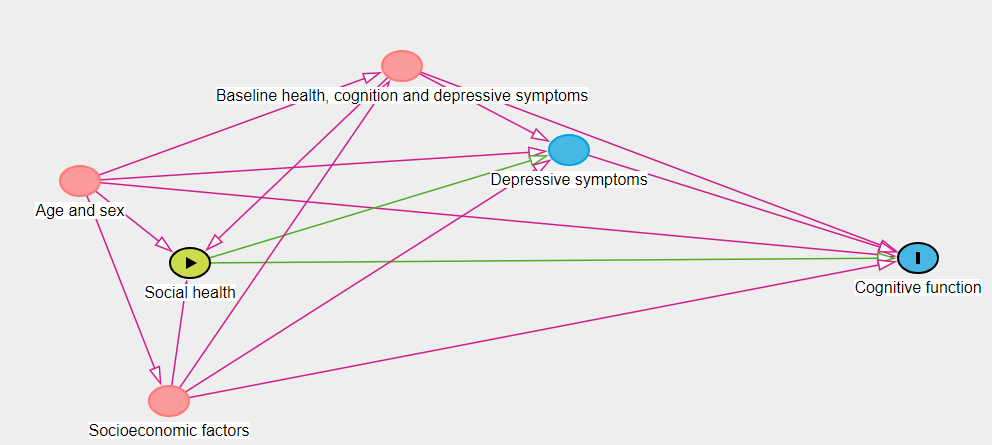

### Figure S3. Directed acyclic graph - inflammation as mediator

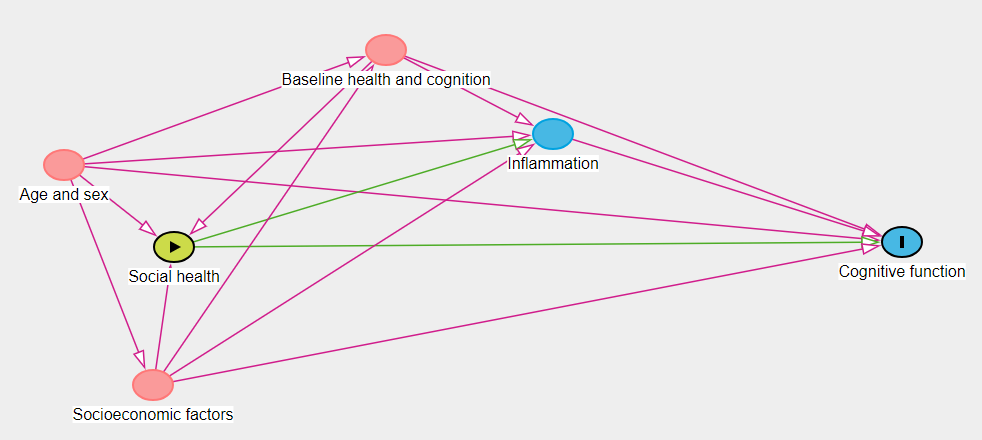

### Figure S4. Four-way decomposition of total effects of social health on cognition (single time point) in SNAC-K - role of depressive symptoms

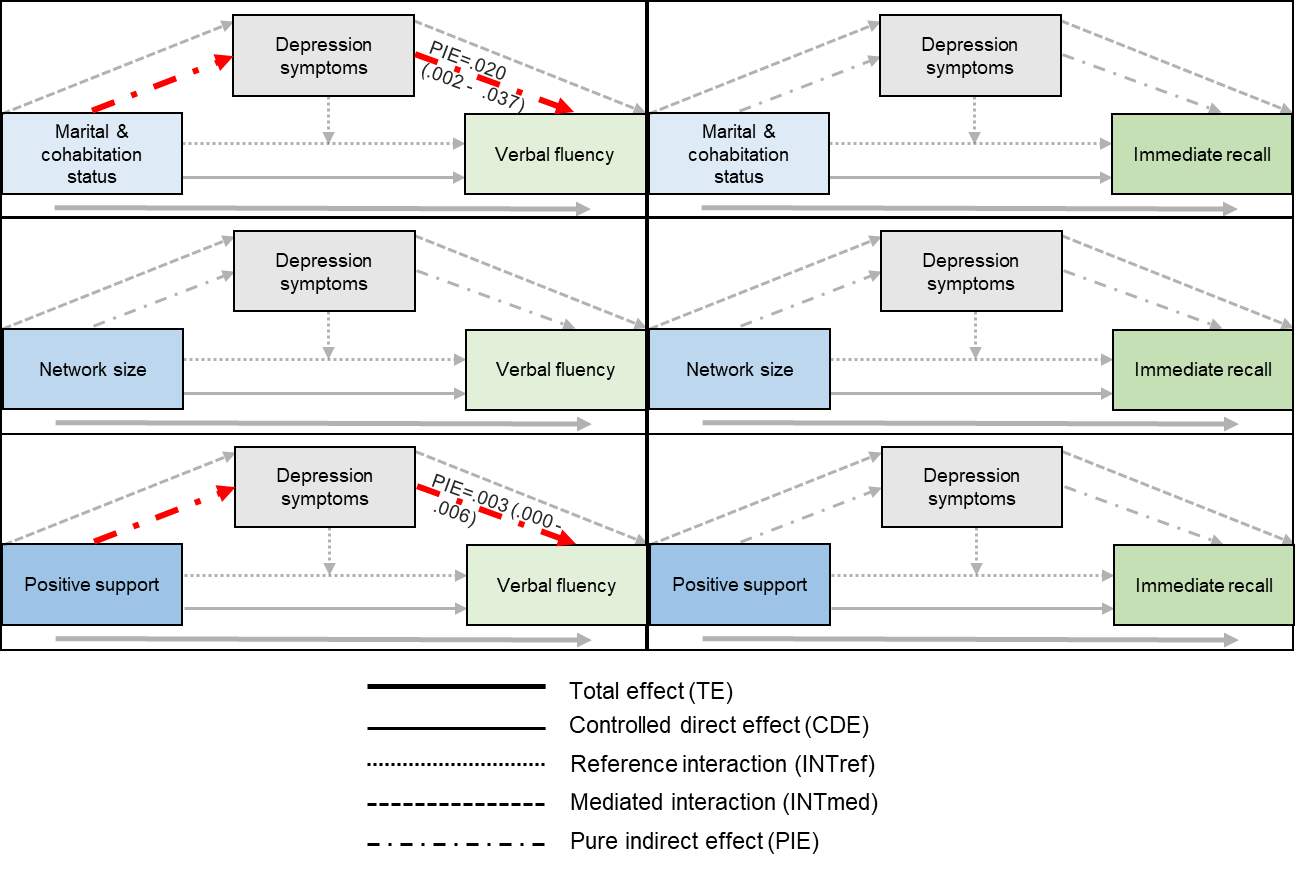

### Figure S5. Four-way decomposition of total effects of social health on cognitive change in SNAC-K - role of depressive symptoms

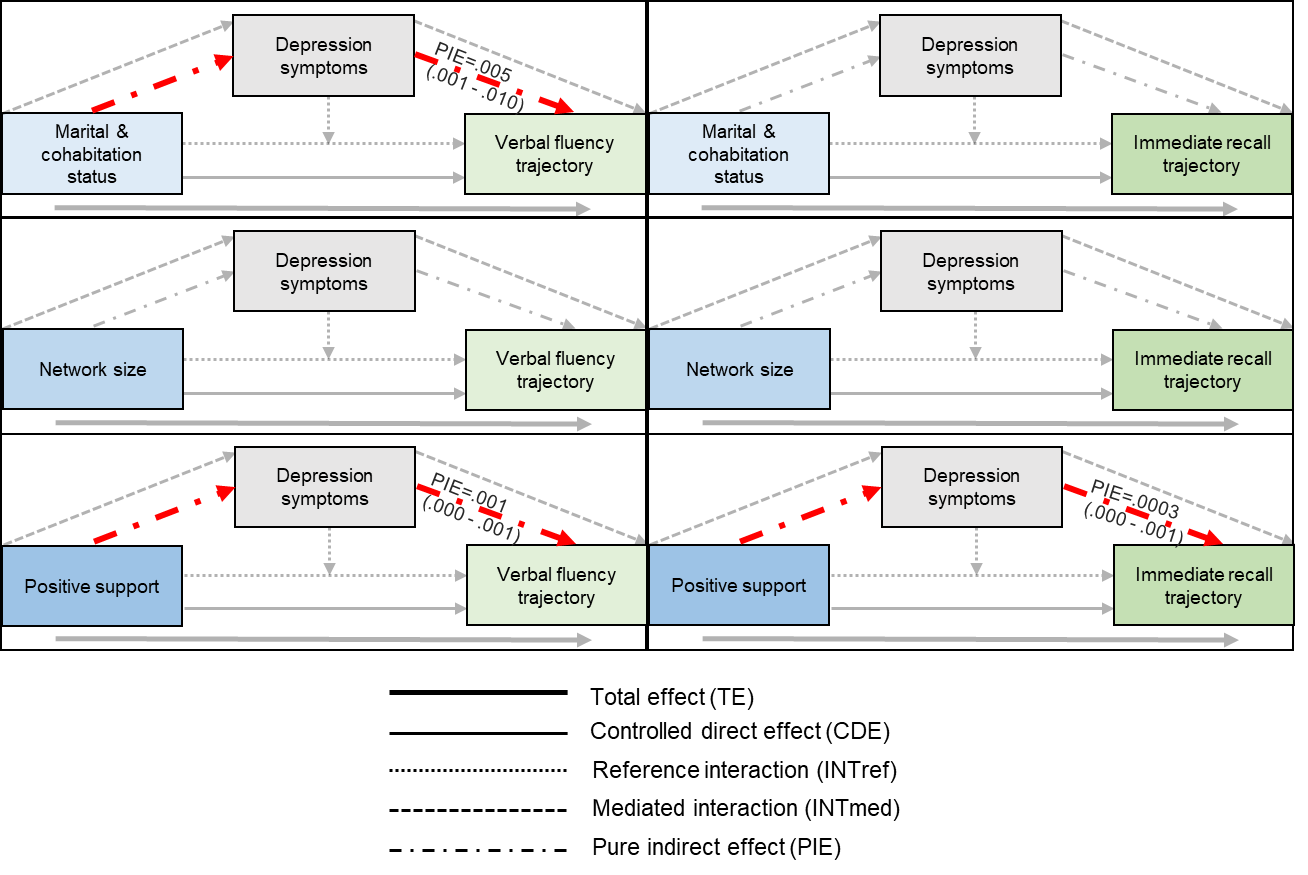

### Supplementary information – Description of social health markers, cognitive outcomes, mediators and covariates in each study

**Description of cognitive tests within each study**

| **Study** | **Description of test** | **N follow-ups** | **Approximate follow-up time (years)** |
| --- | --- | --- | --- |
| **Recall (standardised word recall)** | | | |
| ELSA | Participants were presented orally (using a taped voice) with a word list containing ten words. They were asked to recall as many words as possible both immediately after the reading of the list and after a delay. Different word lists were used to minimise practice effects. We examined associations separately with both immediate and delayed recall. | 7 waves (waves 3-9) | 12 |
| SNAC-K | Participants were presented orally and visually with 6 unrelated nouns (5s pace). Immediately after presentation, participants were given 2 minutes to recall the nouns. Number of words correctly remembered was recorded. | 3 waves (waves 2-4) | 6 |
| **Verbal fluency (standardised verbal fluency)** | | | |
| ELSA | Participants were asked to name as many different animals as possible in one minute. The final score includes the number of animals identified in one minute. | 6 waves (waves 3, 4, 5, 7, 8 and 9) | 12 |
| SNAC-K | Participants were asked to name as many different animals as possible in one minute. The final score includes the number of animals identified in one minute. | 3 waves (waves 2-4) | 6 |

**Structural social relationship variables**

| **Study** | **Question** | **Response** |
| --- | --- | --- |
| **Married or cohabiting (unmarried and alone=0; married or cohabiting=1)** | | |
| ELSA | 1) What is your current legal marital status?  2) Number of people in the household | 1) Single, never married; married; remarried; legally separated; divorced; widowed.  2) Numeric response. |
| SNAC-K | 1) What is your current marital status?  2) Who lives with the participant? | 1) Married (or equivalent); widow/er also in case of civil/common law union); unmarried; divorced.  2) Single; spouse/common-law partner; daughter; son; grandchild; sibling; sister/brother-in-law. |
| **Network size (continuous and categorical - none=0; 1-2 people=1; 3-6=2; ≥6=3)** | | |
| ELSA | For each of the following: children, other family, friends: how many do you have a close relationship with? | Numeric response (*Created a summed score across relationship type).* Overall scores were capped at 30 people. |
| SNAC-K | How many people do you feel you know well and can talk to about most things (e.g., relatives, friends, neighbours, and/or colleagues)? | None; 1-2 people; 3 people; 4-6 people; 7-9 people; 10-15 people; 16-30 people; more than 30 people. |
| **Contact frequency (never to a few times per year=0; about once to twice a month=1; once to three or more times per week=2) *rounded to nearest category for summed scores*))** | | |
| ELSA | For each of the following: children, friends, family:  Q1. How often do you meet up with them?  Q2. How often do you speak on the phone with them?  Q3. How often do you write or email them? | Three or more a week; once or twice a week; one or twice a month; every few months; once or twice a year; less than once a year or never.  We averaged across contact type for each relationship type (children, friends and family) and then averaged across relationship type, rounding to the nearest category to create an overall contact frequency variable.  *Recode to:*  *Less than once a year or never=0; once or twice a year=0; every few months=0; one or twice a month=1; once or twice a week=2; Three or more a week=2.* |
| SNAC-K | For each of the following: parents, children, son or daughter-in-law, grandchildren, siblings, other relative, neighbour, friend:  Q1. How often do you meet them in person?  Q2. How often are you in touch via telephone, letters or email? | Never; less often; quarterly more than once/year; monthly more than six times per year; weekly more than twice per month; daily more than twice per week.  *Recode to:*  *Never or less often=0; quarterly more than once/year=0; monthly more than six times per year=0; weekly more than twice per month=1; daily - more than twice per week=2.* |

**Interactional social relationship variables**

| **Study** | **Question** | **Response** |
| --- | --- | --- |
| **Perceived positive support (numeric score with higher score indicating more positive support)** | | |
| ELSA | For each of the following - spouse/partner, children, immediate family, friends:  1) How much do they really understand the way you feel about things?  2) How much can you rely on them if you have a serious problem?  3) How much can you open up to them if you need to talk about your worries? | A lot; some; a little; not at all.  *Reverse code so not at all=0. Compute mean of each question across relationship type i.e., mean of Q1 for partner, children, family and friend. Then sum across questions.* |
| SNAC-K | 1) Do you feel that you know one or a few people who could give you proper personal/emotional support to manage the stress and troubles of life?  2) Do you know someone with whom you can be yourself, who accepts you for all your good and bad qualities? | Yes, without a doubt; yes, probably; no, probably not; no, not at all.  *Reverse code so not at all=0. Sum.* |
| **Perceived negative support (numeric score with higher scores indicating less negative social support)** | | |
| ELSA | For each of the following - spouse/partner, children, immediate family, friends:  1) How much do they criticise you?  2) How much do they let you down when you are counting on them?  3) How much do they get on your nerves? | A lot; some; a little; not at all.  *Compute mean of each question across relationship type i.e., mean of Q1 for partner, children, family and friend. Then sum across questions.* |
| SNAC-K | Not assessed |  |

**Covariates**

| **Study** | **Description** | **Response** |
| --- | --- | --- |
| **Occupational class (manual=0; non-manual=1)** | | |
| ELSA | Occupational class | Three-class National Statistics – Socioeconomic Classification Scheme:  Managerial and professional occupations; intermediate occupations; semi-routine occupations. |
| SNAC-K | Socioeconomic Index based on type of last/longest-held occupation | Manual; non-manual |
| **Education (lower (lower than secondary education or no education)=0; secondary (secondary education or equivalent)=1; higher (university/other post-secondary)= 2)** | | |
| ELSA | Highest educational attainment | NVQ4/NVQ5/Degree or equiv; Higher ed below degree; NVQ3/GCE A Level equiv; NVQ2/GCE O Level equiv; NVQ1/CSE other grade equiv; Foreign/other; No qualification |
| SNAC-K | Highest educational attainment | Unfinished primary education; Primary school ("folkskola", ca 6 yrs.); Elementary school/Secondary "realskola"/girls' school; High school/Upper secondary/"Gymnasium"; "Folkhögskola" Folk high school/Vocational/Trade school; Education of at least one year after high school graduation; College/University degree |
| **Household wealth (quintiles) (1=lowest quintile, 2=second quintile, 3=third quintile, 4=fourth quintile, 5=highest quintile)** | | |
| ELSA | Quintiles of total (non-pension) wealth | Lowest quintile; second quintile; third quintile; fourth quintile, highest quintile |
| SNAC-K | Not assessed. |  |
| **Basic and instrumental activities of daily living (none=0; at least one=1)** | | |
| ELSA | Because of physical, mental, emotional or memory problems do you have any difficulty with: Walking 100 yards; sitting for about two hours; getting up from a chair after sitting for long periods; climbing several flights of stairs without resting; climbing one flight of stairs without resting; stooping, kneeling or crouching; reaching or extending your arms above shoulder level; pulling or pushing large objects like a living room chair; lifting or carrying weights over 10 pounds, like a heavy bag; picking up a 5p coin from a table; dressing, walking across the room; showering or bathing; preparing a hot meal; eating, such as cutting up food; getting in or out of bed; using the toilet, including getting up or down; using a map to figure out how to get around in a strange place; shopping for groceries; making telephone calls; taking medications; doing work around the house or garden; managing money, such as paying bills and keeping track of expenses. | Yes/no option for each item |
| SNAC-K | Participants were asked whether they could: independently manage their daily activities (e.g. cooking, cleaning, and running errands); buy grocery; prepare a meal; manage household chores (light and heavy); manage laundry; manage household economy; use the telephone; use public transports; drive a car (if participant’s had access to a car). |  |
| **Vascular-related health conditions (0=none; 1=at least one)** | | |
| ELSA | Self-report doctor diagnosed: angina, heart attack/myocardial infarction, congestive heart failure, heart murmur, abnormal heart rhythm, diabetes or high blood sugar, stroke/cerebral vascular disease, other heart trouble. | Yes/no option for each diagnosis |
| SNAC-K | TYPE 2 DIABETES: self-reported medical history, glucose-lowering medication use, medical records from the NPR (ICD-10 code E11), or glycated haemoglobin ≥6.5%.  HEART DISEASES (CVD) including: 1) atrial fibrillation (ICD-10 code I48 and/or discrete P-wave undetectable and irregular ventricular rate), 2) bradycardias and conduction diseases (presence of a cardiac pacemaker and ICD-10 codes I441-I443, I453, I455, Z950), 3) ischemic heart disease (ICD-10 codes I20-I22, I24-I25, Z951,Z955, and/or use of organic nitrates [Anatomical Therapeutic Chemical; ATC, code C01DA] or ranolazine [ATC code C01EB18]), 4) cardiac valve disease (ICD-10 codes I05-I08, I091, I098, I34-I38, I390-I394, Q22-Q23, Z952-Z954), and 5) heart failure (ICD-10 codes I110, I130, I132, I27, I280, I42-I43, I50, I515, I517, I528, Z941, Z943).  CEREBROVASCULAR DISEASES included stroke/TIA and other cerebrovascular syndromes. Diseases were identified according to the ICD-10 codes G45-G46, I60-I64, I67, and I69. |  |
| **Depressive symptoms (higher scores indicate more depressive symptoms. Also created binary variable using cut-off score indicating high vs low symptoms)** | | |
| ELSA | 8-item Center for Epidemiologic Studies Depression Scale (CES-D). | The CES-D is widely used to identify people at higher risk of depression in population-based studies. Items provide information on negative affect and somatic complaints experienced in the past week. Dichotomous responses to each item (yes/no) result in a total score ranging from 0 (no symptoms) to 8 (all eight symptoms). We reverse coded items so that higher scores indicated more depressive symptoms. All items were summed. We examined depressive symptoms both as continuous, and as a binary variable using a score of four or above to index clinically significant depression as recommended (Steffick et al., 2000), which corresponds with the cut-off point of 16 on the 20-item CES-D. |
| SNAC-K | Montgomery-Åsberg Depression Rating Scale (MADRS) from Comprehensive Psychopathological Rating Scale | 10 items, including low mood, inner tension, sleep disturbances, change in appetite, pessimistic thoughts, lassitude, inability to feel, apparent sadness, suicidal thoughts and concentration difficulties. Continuous (sum of symptoms; range 0–60); >6 is clinically relevant |

| **Physical activity (0=highly active (vigorous activity at least once a week), 1=moderately active (moderate activity at least once a week), 2=inactive (no moderate or vigorous activity on a weekly basis)** | | |
| --- | --- | --- |
| ELSA | Frequency of participation in vigorous, moderate and light exercise per week. | 1=more than once a week, 2=once a week, 3= one to three times a month, 4=hardly ever, or never |
| SNAC-K | Frequency of participation in light, moderate, intense exercise per month in light and/or moderate/intense | 1=inadequate (never, <2–3 times per week) 2= health-enhancing (light exercise several times per week or every day) 3= fitness-enhancing (moderate/intense exercise several times per week or every day) |

| **Smoking (0=never, 1=ex-smoker, 2=current smoker)** | | |
| --- | --- | --- |
| ELSA | Participants asked about smoking status | 0=never smoked, 1=ex smoker - occasional, 2= ex smoker - regular, 3= ex smoker - don't know frequency, 4=current smoker  *Recode to:*  *Never smoked=0; ex-smoker - occasional=1; ex-smoker - regular=1; ex-smoker - don’t know frequency=1; current smoker=2.* |
| SNAC-K | Participants asked about smoking status Never/former/current | |

| **Alcohol intake (ELSA 0=special occasions, or once or twice a month, 1=around weekly, 2=around daily, 3=not at all; SNAC-K** **0=no or occasional, 1=light to moderate, 2=heavy)** | | |
| --- | --- | --- |
| ELSA | In the past 12 months have you taken an alcoholic drink (twice a day or more, daily or almost daily, once or twice a week, once or twice a month, special occasions only, or not at all) | 1=twice a day or more, 2=daily or almost daily, 3=once or twice a week, 4=once or twice a month, 5=special occasions only, 6=or not at all  *Recode to:  Special occasions=0, once or twice a month=0, once or twice a week=1, daily or almost daily=2, twice a day or more=2, Not at all=3* |
| SNAC-K | Drinking habits in a typical week No or occasional, light to moderate (1–14  drinks/week for men [M] and 1–7  drinks/week for women [W]), or heavy  (≥15 drinks/week [M], and ≥8 | |
|  | drinks/week [W]). | |

**Mediators**

| **Study** | **Description** | **Response** |
| --- | --- | --- |
| **Depressive symptoms (Continuous - higher score indicates more depressive symptoms. Binary - cut-off score indicating high vs low symptoms)** | | |
| ELSA | 8-item Center for Epidemiologic Studies Depression Scale. | Yes/no for each item. Reverse code items so that higher score indicated more depressive symptoms. All items summed. Cut-off for binary outcome: 4. |
| SNAC-K | Montgomery-Åsberg Depression Rating Scale (MADRS) | 10 items, including low mood, inner tension, sleep disturbances, change in appetite, pessimistic thoughts, lassitude, inability to feel, apparent sadness, suicidal thoughts and concentration difficulties. Continuous (sum of symptoms; range 0–60); >6 is clinically relevant |
| **CRP (continuous – Mg/l)** | | |
| ELSA | Blood samples collected during nurse visit and analysed in the Royal Victoria Infirmary (Newcastle-upon-Tyne, UK). Circulating high-sensitivity CRP was assessed using the N Latex high sensitivity CRP mono immunoassay on the Behring Nephelometer II Analyzer. The limit of detection was 0.17 mg/l and the coefficient of variation was less than 6% for this assay. | Mg/l. To account for skewed distributions, CRP was log-transformed. Blood samples taken from willing ELSA core members, except those who had a clotting or bleeding disorder, had ever had a fit, were not willing to give written consent, or were currently on anticoagulant drugs. Fasting blood samples taken whenever possible, although respondents aged over 80 years, those who seemed frail, or where the nurse was concerned about their health, were not asked to fast. Subjects considered to have fasted if they had not had food or drink except water for a minimum of 5 hours before the blood test. |
| SNAC-K | Non-fasting venous blood samples centrifuged for 10 minutes at 3,000 runs per minute, and kept at 2°C. CRP serum concentrations were analysed at Karolinska University Hospital, Stockholm, within 3 hours from sampling, through an enzyme-linked immunosorbent assays with a lower detection limit of 1 mg/L. | Mg/l with cut-offs for 0-5; 6-10, 11-20 20+ |
| **Fibrinogen (continuous – g/l)** | | |
| ELSA | Blood samples collected during nurse visit and analysed in the Royal Victoria Infirmary (Newcastle-upon-Tyne, UK). Fibrinogen levels were assessed using the Organon Teknika MDA 180 analyser, using a modification of the Clauss thrombin clotting method, with a coefficient of variation of less than 10%. | g/l. Fasting blood sample process as described above in relation to CRP. |
| SNAC-K | Not assessed. |  |
